# Modeling COVID-19 for lifting non-pharmaceutical interventions

**DOI:** 10.1101/2020.07.02.20145052

**Authors:** Matt Koehler, David M. Slater, Garry Jacyna, James R. Thompson

## Abstract

As a result of the COVID-19 worldwide pandemic, the United States instituted various non-pharmaceutical interventions (NPIs) in an effort to the slow the spread of the disease. Although necessary for public safety, these NPIs can also have deleterious effects on the economy of a nation. State and federal leaders need tools that provide insight into which combination of NPIs will have the greatest impact on slowing the disease and at what point in time it is reasonably safe to start lifting these restrictions to everyday life. In the present work, we outline a modeling process that incorporates the parameters of the disease, the effects of NPIs, and the characteristics of individual communities to offer insight into when and to what degree certain NPIs should be instituted or lifted based on the progression of a given outbreak of COVID-19.

## Introduction

In December of 2019, a cluster of pneumonia cases of unknown origin were identified in Wuhan, China. An investigation into the cases commenced in early January 2020 that led to the discovery of a novel coronavirus now designated SARS-CoV-2. The virus causes an infectious disease now known as Coronavirus Disease 2019 or COVID-19. Common symptoms of COVID-19 include shortness of breath, fever, dry cough, fatigue, and respiratory distress.

On January 10^th^, 2020 there were 41 confirmed cases of COVID-19 in China [1]. By January 20^th^, *The Guardian* reported that the Chinese National Health Commission had confirmed human-to-human transmission of COVID-19 and the number of reported cases had more than tripled to 139 [2]. That same day, Chinese authorities started implementing non-pharmaceutical interventions (NPIs) such as social distancing (staying at least six feet from other humans) and limiting travel. On March 11^th^, 2020, the World Health Organization declared a global pandemic. By mid-June 2020 there were over 8.8 million cases worldwide and global deaths exceeded 462,000 according to Johns Hopkins University [3]. COVID-19 cases have been confirmed in 188 countries around the world and all 50 states of the U.S.

As of June 2020, there was no vaccine or known antiviral medication effective against COVID-19 [4]. Treatment involves addressing the symptoms as the virus runs its course. Therefore, most countries have implemented NPIs in an effort to prevent the spread of COVID-19. These interventions usually involve restricting the movements of the population. The U.S. has employed school closures, shelter-in-place orders, business closures, limits on or banning of public gatherings, closing public spaces such as parks, and social distancing orders that require people to stay at least six feet from each other. In addition, a combination of testing and tracing anyone who came in contact with a positive case – referred to as contact tracing – and requiring them to self-quarantine for approximately 14 days is another tactic for limiting the spread.

The restrictions to daily life have driven the world into one of the largest recessions in history [5]. Goldman Sachs has warned that the U.S. Gross Domestic Product (GDP) could contract as much as 29% in the second quarter and the U.S. unemployment rate rose from 3.6% in January 2020 to 14.7% in April 2020 [6]. The economic recession creates an urgency to lift the NPIs and population restrictions that is in direct conflict with the desire to stop the spread of disease and keep the population healthy. State and federal leaders therefore need tools that can provide insight into the risk to populations of lifting NPIs before the disease is completely wiped out or a vaccine is developed.

Empirical studies have shown that the same disease can exhibit heterogeneous transmission characteristics as it spreads to different communities [7]. This fact has led researchers to look for ways to more realistically model the contact structure of a given population. In a separate paper we derived a mathematical framework that results in a closed-form analytical solution using percolation theory and mean-field theory. The analytical model uses the degree distribution of a contact network and the parameters of the disease to estimate properties of an outbreak. A limitation of the analytical framework is that it assumes *a priori* knowledge of the contact network degree distribution. In the current work, we establish a procedure for inferring the degree distribution of a given county in the U.S. from Census data [8]. This provides a pre-pandemic community structure. We then constructed an agent-based model (ABM) that incorporates the daily behavior of individuals in a given community, along with the constraints to that behavior imposed by NPIs. The ABM ingests the pre-pandemic community network inferred from Census data to drive social contact when no NPIs are implemented. The post-pandemic contact structure emerges as a function of compliance with NPIs and we are able to assess differences in the spread of disease as a function of both contact network degree distribution and the impact that a given set of NPIs has on the network structure.

To illustrate the utility of our approach we model the 24 county-equivalents of Maryland in an experiment that asks *what is the impact of a given percent of the population being tested and a given level of participation in contact tracing if NPIs are lifted 70 days after the onset of the pandemic?* We show that different strategies can have the same impact on disease progression depending on the contact structure of each county. This underscores the importance of the modeling capability because it facilitates a location-based phased approach for lifting NPIs and instituting test and trace strategies. It also serves to underscore the importance of incorporating community structure into the analysis. Seven of the 24 county-equivalents of Maryland show a propensity for much larger outbreaks of COVID-19 than the rest of the state. We use the previously derived analytical model and empirically collected case data to validate the results of the ABM. The ABM combined with the analytical model allows decision-makers to determine which factors are most important in classifying the risk of large outbreaks in different regions and facilitates the customization of NPI strategies.

## Materials and methods

### The Analytical Model

Throughout this section we use precise definitions of certain properties of epidemiological models based on Meyers et al., Bettencourt et al. and Chowell et al. [7, 9, 10].

- **Transmissibility** *T* is the average probability that an infectious individual will transmit the disease to a susceptible individual with whom they have contact.
- **Critical Transmissibility** *T*_*c*_ is the minimum transmissibility required for an outbreak to become a pandemic. 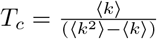 where ⟨*k*⟩ and ⟨*k*^2^⟩ are the mean and variance of the degree distribution of the contact network.
- **Basic Reproduction Number** *R*_0_ is the expected number of cases directly generated by one case in the population of susceptible individuals. It can be shown to be equal to the ratio of transmissibility to the critical transmissibility 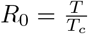.
- **The Instantaneous Reproduction Number** 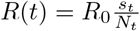 where *s*_*t*_ is the number of susceptible individuals at time *t* and *N*_*t*_ is the total population.

Note that some studies refer to an effective reproductive number *R*_*e*_. In the definitions above, *R*_0_ depends on the degree distribution so there is no need to make this distinction. *R*_0_ and *R*_*e*_ can be thought of as interchangeable terms.

Given the transmission rate *r* between vertex *i* and vertex *j* of a network graph and the infection time *τ* and assuming that *r* and *τ* are independent random variables, the average transmissibility is:

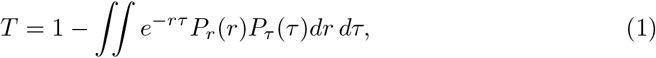

where *P*_*r*_(*r*) and *P*_*τ*_ (*τ*) are the respective probability density functions (pdfs). For simplicity, it is assumed that *P*_*r*_(*r*) = *δ*(*r − r*_0_) and *P*_*τ*_ (*τ*) = *δ*(*τ − τ*_0_) so that 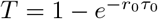.

For a randomly chosen vertex, let *p*_*k*_ denote the probability that this vertex has *k* edges. Then *G*_0_(*x*) is the generating function for the degree distribution of this vertex:

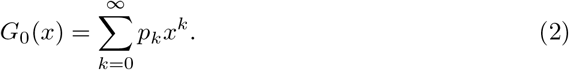

An important result by Feld is that the degree distribution of the first neighbor of a vertex is not the same as the degree distribution of vertices as a whole [11]. There is a higher chance that an edge will be connected to a vertex of high degree, in fact, in direct proportion to its degree. Let *q*_*k*_ denote the degree distribution of a vertex at the end of a randomly chosen edge. Then:

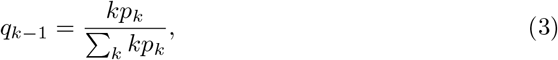

excluding the randomly-chosen edge. The corresponding generating function for this distribution is:

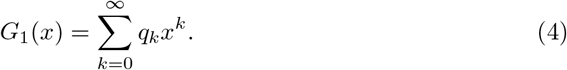

The generating functions *G*_0_(*x*) and *G*_1_(*x*) are related. Let *z* = Σ_*k*_ *kp*_*k*_ and using Eq. (3):

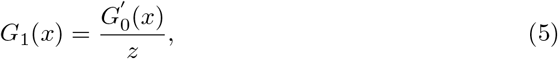

where 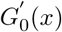 is the derivative of *G*_0_(*x*).

Using bond percolation and mean-field theory, Newman [12, 13] showed that for small outbreaks (clusters) when *T < T*_*c*_ we can combine Eq. 1 with the generating functions and then compute the average cluster size ⟨*c*⟩ as:

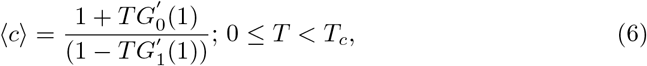

where the angle brackets indicate the expected value. In the case of a pandemic where *T > T*_*c*_ we define *H*_1_(*x*; *T*) as the generating function of the size of a cluster at the end of a randomly chosen edge. We can then derive an iterative approach for calculating the probability *P* of a pandemic. Let *v* = *H*_1_(1; *T*), then the fixed point problem becomes:

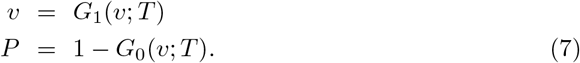

We expanded on this formulation to incorporate the NPIs of uniform and directed social distancing. For social distancing, let *b*_*k*_ denote the probability that a vertex of degree *k* is present. Define the generating function *F*_0_(*x*) as:

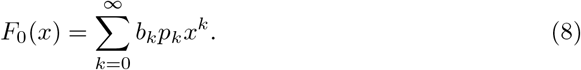

Similarly, 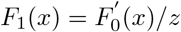. This is a generalization of the generating functions *G*_0_(*x*) and *G*_1_(*x*). Since *F*_0_(1) *≠* 1 and *F*_1_(1) *≠* 1, 1 *− F*_0_(1) is the probability that a randomly chosen vertex has no active edges. In the case of uniform social distancing, vertices are chosen to sequester at random so *b*_*k*_ = *b*.

In the case of directed social distancing, it is assumed that:

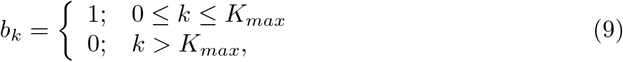

where all individuals are distanced with contact degree greater than *K*_*max*_. This implies that:

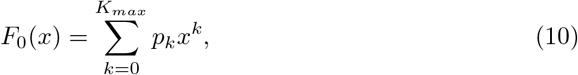

To compute the size ⟨*c*⟩ of small outbreaks (*T < T*_*c*_) the generating functions 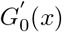 and 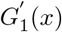 are simply replaced with 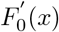 and 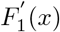 respectively. For large pandemics, the derivation is similar and the resulting fixed point problem is:

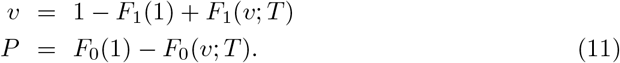

Incorporating these generating functions into a time-dependent model is straight forward. If we define *v*(*t*) as the average probability of a neighbor (connected via an edge) being infected and *w*(*t*) as the average probability of recovery, then it can be shown that the time-dependent spread of disease on a network with generating distribution *G*_1_(*x*) and no NPIs is defined by:

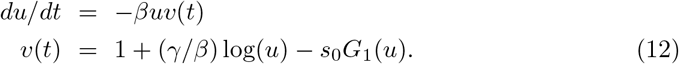

where *u*(*t*) = exp(*− βw*(*t*)*/γ*), *β* is the contact rate, *γ* is the recovery rate, and *s*_0_ is the proportion of initially susceptible individuals [14].

To model the time-dependent spread when NPIs are implemented and then lifted, we can define different intervals of time where *v*(*t*) is modified to reflect the decrease in potentially infected neighbors. In the case of uniform social distancing we replace *v*(*t*) with:

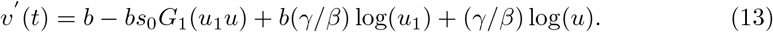

were *b* is the fraction of the population that is not sequestered. In the case of testing and contact tracing, the model becomes more complicated, requiring another ordinary differential equation *v*‴ (*t*) to replace *v*(*t*) such that:

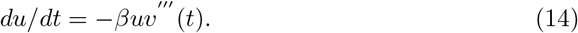

The derivation of the full model is beyond the scope of the current work, so we refer the interested reader to our companion paper.

### Inferring Network Structure

The Analytical Model requires *a priori* knowledge of the social contact network and its degree distribution in order to assess the risk of an outbreak. We are thus faced with the need to infer the network from data collected on a given population. Fortunately, the U.S. Census conducts an extensive annual survey known as the American Community Survey (ACS) [8]. The ACS provides county level information on characteristics of the population such as households, household size, school enrollment, occupation, and age distribution of the population. Meyers et al. outlined a process for taking similar data (their study focused on Vancouver, Canada) and inferring a network structure based on a few simplifying assumptions [7]. Here we design a similar algorithm using the ACS data to create county-level social contact networks that are likely to be representative of human-to-human contact *before* COVID-19 and the associated NPIs.

Meyers et al. assumed that individuals living in the same household would have physical contact with probability *p*_*hh*_ = 1.0. That is, every vertex in a household has an edge with every other vertex in the same household. Once the household subgraphs are created, the population is then assigned to schools based on their age group and school enrollment data and workplaces based on occupation data. Individuals who attend the same school have a physical contact with probability *p*_*s*_ and those who work together have contact with probability *p*_*w*_. Finally, people have friends, go to restaurants and stores, and otherwise interact socially. These public contacts occur with probability *p*_*pub*_ and can involve any two individuals in the community. The full algorithmic statement is as follows:

**Inputs:** U.S. Census ACS data, state, county, *N* = total desired vertices

1. Create an array of *N* total vertices distributed according to age in the given county
2. Randomly choose *a*_65_ vertices from those over 65 years of age to live alone where *a*_65_ is taken from the ACS field for Householder alone over 65
3. Randomly choose *a* vertices from those between 18 and 65 years of age to live alone where *a* is taken from the ACS field Householder alone under 65
4. From the remaining vertices over 18 years of age, randomly choose *mc* pairs of nodes where *mc* is taken from the ACS field Married Couples
5. From the remaining vertices under 18 years of age, randomly assign *mc*_18_ vertices to a married couple where *mc*_18_ is taken from the ACS field Married Couple with one or more children
6. From the remaining vertices under 18 years of age, randomly assign *s* vertices to a single adult over 18 years where *s* is taken from the ACS field Single with one or more children
7. Determine the remaining households not created and randomly group remaining vertices into those households by assigning edges so that average household size equals the corresponding ACS field
8. For each enrollment population *n*_*i*_ taken from the five ACS school enrollment fields, randomly choose 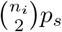 vertices and create one edge for each vertex with a second randomly chosen set of 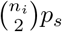 vertices taken from the appropriate age group of vertices
9. For each occupation population *n*_*i*_ taken from the 13 ACS occupation fields, randomly choose 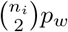 vertices and create one edge for each vertex with a second randomly chosen set of 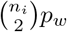 vertices taken from the set of vertices over the age of 16 and assigned to that occupation
10. Randomly choose 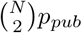 vertices from the set of all vertices and create an edge with a second randomly chosen set of 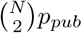 vertices taken from the set of all vertices ensuring no duplicate edges or self-loops

**Return:** social contact network *G* = *{V, E}* where *V* is the set of vertices and *E* is the set of edges.

The resulting progression from households to schools to work to public is illustrated through the four stages of construction in Fig. 1.

**Fig 1.**
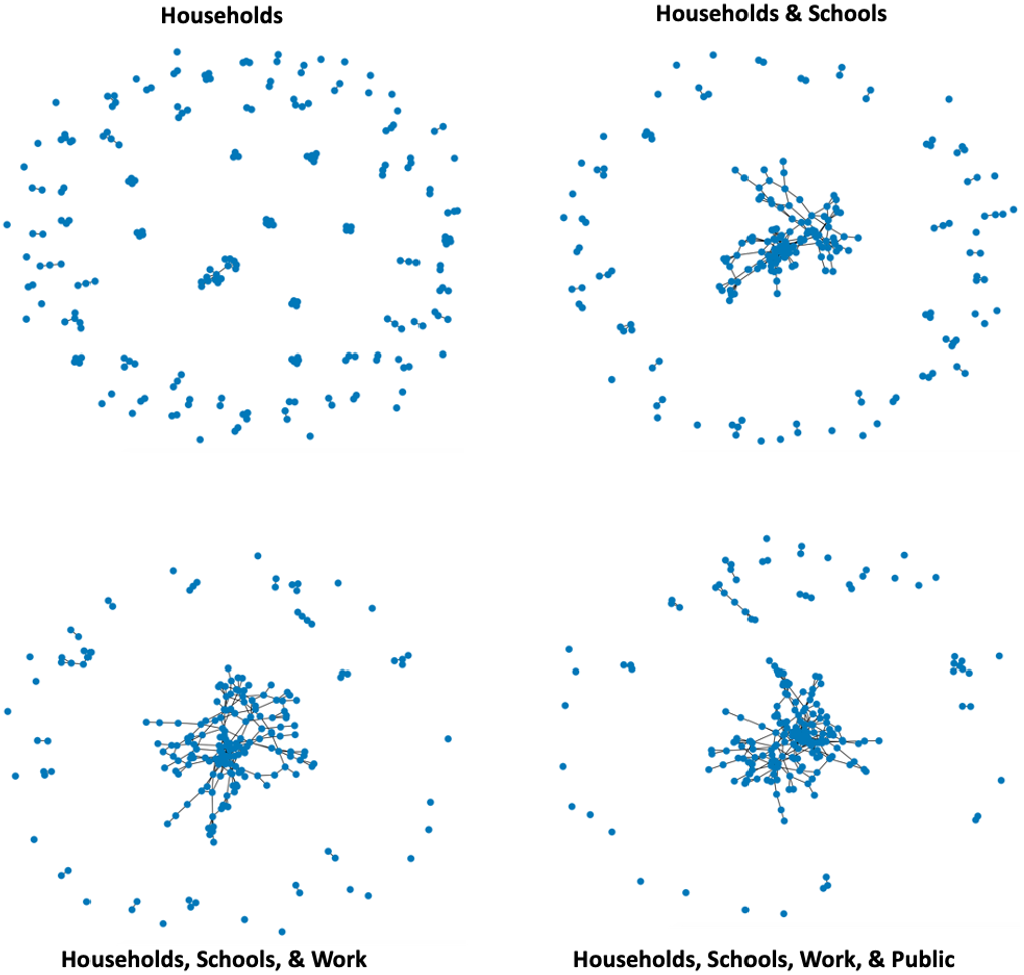
Stages of Network Construction from Census Data

### The Agent-based Model

Disease propagation is a complex system [15]. The dynamics of a pandemic is a function of the biological characteristics of the disease, the biologic response of those infected, and the behaviors of those in the area affected by the disease. In this way a pandemic is an emergent phenomena of the social systems within which it is embedded [16].

Emergent phenomena are a characteristic of complex systems [17, 18], systems made up of many interacting, heterogeneous entities embedded in a relevant space. In these types of systems the interactions among entities are as important to the overall dynamics as the characteristics of the entities themselves. Therefore, a strict reductionist approach will only provide partial insights into the system dynamics. For example, a detailed biological analysis of a virus will only provide limited insights into how an eventual pandemic may unfold. Regardless of the contagiousness of a virus, if all members of a society maintain strict social distancing, the disease cannot spread. Given that this is a complex system, an efficient way to discover its future state is to simulate it [19]. Finally, the most natural way to express these systems as a simulation is to do so as an agent-based model (ABM), [20, 21].

Although our analysis is premised on inferred contact networks, we felt the importance of exploring the dynamics associated with changes in behavior warranted the inclusion of an ABM so one could more easily explore the impact of non-pharmaceutical interventions (NPIs) and the rate of compliance. Furthermore, creating this ABM allowed us to explore the basic reproduction number *R*_0_ of the pandemic as an emergent phenomena and one that is potentially unique to each infected individual. In this way such things as super spreaders do not need to be explicitly included as an exogenous “shock” but rather they emerge based upon the behavior of the individual members of the society.

#### Overview of the ABM

The ABM is implemented in NetLogo [22]. The model is instantiated first with an input file that defines the inferred contact network of a U.S. county scaled to be approximately ten thousand individuals. These contact graphs are generated using U.S. Census data and the algorithm described in the section Inferring Network Structure.

The environment size is scaled to approximate the population density of the county in question. First, all members of the simulated society are created. Second, school, work, home, and public contact networks are created. The physical space is created next. For each connected component of the aforementioned graphs, citizens are assigned to a school, work site, public venue, or home. All disconnected agents in the home graph are assigned a home individually, as well. The locations are then placed in the environment. They can be mixed (as one might see in an urban area where individuals live, work, shop, and learn in very close proximity) or home sites can be separated from work, school, and public venues (as one might see in rural or suburban counties). A user specified number of citizens are then infected. The last step in simulation initialization is to remove all links, at this point all contact is driven by behavioral dynamics.

At run time each agent spends the evening at home and the day at work, school, or a public venue if they have one and these non-home sites are open. All contagious agents attempt to spread the disease each time step. As discussed elsewhere how well the disease propagates in the society is a function of individual mitigation steps and NPI imposed.

The available NPIs include closing schools, closing work places, closing public venues, imposing social distancing requirements (i.e., stay home orders), and isolating individuals. Individual mitigation steps can also be modeled using the probabilities associated with disease spread. Testing may also be performed within the simulated society. Two different testing strategies can be employed. The first is random testing of non-hospitalized individuals. The second distributes the tests across symptomatic and asymptomatic individuals. Once tested or found to have a positive test, individuals can be isolated for some period of time (e.g., three days while waiting for results or two weeks if found to be positive). Last we incorporated contact tracing that is triggered by either a positive test or a hospital visit. Agents who have opted-in to the contact tracing are informed to self-isolate if they came in contact with an infected agent who subsequently tested positive or went to the hospital.

The simulation ends when all individuals are no longer contagious (all individuals are in the susceptible, deceased, or recovered states) or when a preset time step limit is reached.

#### Disease Propagation

The disease progression model was designed to follow the dynamics of COVID-19 as they were understood around March of 2020. Similar to a classic SEIR model, the agents move through discrete states as the disease progresses within them. The disease states include Susceptible, Exposed, Mild, Severe, Critical, Deceased, and Recovered. All agents are instantiated in a state of Susceptible. Immediately prior to runtime a user specified number of agents are set to the Exposed state.

In order to increase flexibility and respond to changes in the understood dynamics of COVID-19 disease progression, the simulation uses default values that can be overridden. The default dynamics are outlined in Tables 1 and 2.

**Table 1.** COVID-19 Progression Parameters.

| Time Period | Duration |
| --- | --- |
| Exposed Period | 5 days |
| Mild Period | 6 days |
| Severe Period | 4 days |
| Critical Period | 10 days |

**Table 2.**
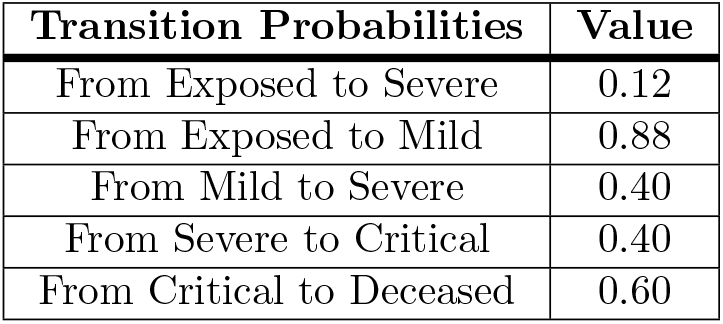
COVID-19 Transmission Parameters.

| Transition Probabilities | Value |
| --- | --- |
| From Exposed to Severe | 0.12 |
| From Exposed to Mild | 0.88 |
| From Mild to Severe | 0.40 |
| From Severe to Critical | 0.40 |
| From Critical to Deceased | 0.60 |

If, at the time of transition from one state to the next, the probability draw is not successful, the agent recovers from the disease. For example, once successfully Exposed the agent will move to either the Mild (0.88 chance) or Severe (0.12 chance) state. If the agent transitions to the Severe state, then after 4 days they will transition to either the Critical state (0.40 chance) or the Recovered state (0.60 chance). If on the other hand, the agent transitions to the Mild state from the Exposed state, after 6 days they will either transition to Severe state (0.40 chance) or the Recovered state (.60 chance). Once in the Recovered state agents are no longer able to contract the disease. Finally, agents in the Critical state are assumed to need breathing assistance and significant medical support; therefore, if the agent is unable to go to the hospital within 3 days of becoming critical they will die with probability 1.0.

Agents are contagious when they are in the Mild, Severe, or Critical states. It is assumed that they are asymptomatic and not contagious in the Exposed state.

#### Transmission of Disease

When agents are collocated, there is some chance that the disease will be transferred from one agent to another. At each time step agents that are contagious (those in the Mild, Severe, or Critical states) will look for another agent on the same patch (small piece of the environment). If there is at least one other agent on the patch contagious agents will attempt to pass the virus to one other agent. Successful passing of the virus is a function of 1) a user specified probability of successfully getting the virus on the other agent (e.g., is the infected agent wearing a mask or not), 2) a user specified mitigation probability (e.g., did the potentially infected agent wash their hands a lot and maintaining a distance of 6 feet from others), and 3) the health status of the target agent (if they are already sick, they will not get sicker).

#### Contact Tracing and Testing Formulation

When citizens are initialized they ‘decide’ to participate in contact tracing (opt-in) or not. This is done via a random draw against a user-specified parameter. A uniform pseudo-random number is generated and if that number is less than the user-specified threshold the citizen participates in contact tracing. Once set, this is held constant for the duration of the run.

Contact tracing can be triggered in two basic ways. The first trigger for contact tracing is when a symptomatic individual arrives at the hospital. The second is via testing. If an individual tests positive and that tested individual has opted-in to the contact tracing program, then contact tracing from that individual will commence.

At runtime (with the appropriate runtime settings) each citizen collects data on the other citizens it comes into contact with. A time step is currently defined as one 12-hour period and a patch (the smallest unit of the landscape in the model) is a square with each side 0.1 km in length. Given this scale, we assume citizens that are co-located on a patch are likely enough to be in significant contact with each other as to warrant being considered in the contract trace. NOTE: all agent IDs are collected whether or not they have opted-in to the contact tracing program, this is done so we can produce a contact network to analyze separately. This decision also improves runtime efficiency, as it is far faster to query for the agents on a patch than to query for agents within a certain distance.

At the beginning of each time step, agents update their health status and then collect contact data in a first-in-first-out queue that is 28 elements long (14 days). For this discussion we will assume the tracing is precipitated by testing a segment of the population for COVID-19. All the citizens who tested positive are passed to the trace procedure and stored in a local variable. If that citizen has opted-in to the contact tracing program, their contact list is then added to the local variable.

Once all contacts have been gathered from citizens who opted-in to the contact tracing program, they are consolidated into a single set. Each citizen contained in the set who have opted-in to the contact tracing program are then told to begin isolation.

#### Testing

In broad terms, testing is accomplished in the simulation in one of two ways. Version 1 testing corresponds to random testing of all individuals outside of the hospital. In this case a user-specified number of citizens are randomly chosen to be tested whenever testing takes place. For version 2 of testing, at a given point in time a number of tests are made available. This number is divided between symptomatic and asymptomatic citizens. A set of citizens equal to or less than the number of allocated tests is then randomly chosen and tested. Testing accuracy includes a user-defined false negative rate, but false-positives are not modeled. This essentially means there is a 100% chance that someone who received a positive test has COVID-19, but a citizen receiving a negative test result is not guaranteed to be free of the disease.

Settings for testing can all be manipulated during runtime so there are no initialization components. Users can choose which version of testing to use and set the percent of the population to test. Alternatively, a user can specify an interval of testing that starts and stops at specific time steps. The way the population is tested is random with replacement. This means, for example, if the interval is set to 1 and 50% of the population is to be tested, then on average half the population will be tested once a day, but a specific citizen might be tested twice or not at all.

First, we calculate the number of tests available by multiplying the number of citizens by percent to be tested parameter. This value is rounded to the nearest whole number. Next we calculate how many of the total number are to be used on symptomatic citizens. That number is created by multiplying the total number of tests by the user-specified split value. Then we calculate the number of tests to be used on asymptomatic citizens by subtracting the total symptomatic from total number of tests. We then create two sets of citizens to test, one for symptomatic and one for asymptomatic. Note there is no guarantee that there will be enough citizens to satisfy the number of tests available. Symptomatic citizens are defined as those with “Mild,” “Severe,” or “Critical” COVID-19 disease progression. Citizens with “Healthy,” “Exposed,” or “Recovered” are considered asymptomatic. If there are extra tests those tests are not used. The next step in testing is to determine the citizens who tested positive. For the symptomatic group, since the only disease in the model is COVID-19, that calculation is straightforward. We take the total number of individuals being tested and multiply that by the accuracy of the test. That gives us the symptomatic citizens with positive tests. The asymptomatic group is slightly different. That group contains citizens who do not have the disease as well as those that do because we model false negatives with test accuracy. To deal with this we need to grab only those individuals who actually have COVID and then apply the test accuracy to that group to get the number of citizens who are asymptomatic and will receive a positive test.

Finally, we need to isolate these citizens. All citizens who are tested are placed in isolation for the duration of the testing period (a user-defined time captured). Those who test positive are placed in isolation for two weeks and, if applicable, contact tracing is done.

#### Initialization and Running of the Agent-Based Model

Initialization of the model is centered around the inferred contact network described previously. The input file contains two basic parts: 1) a list of agents, and 2) an edge list that specifies source and sink agents and edge type. Edge types can be one of the following: home, work, school, or public.

First, we set the size of the world as a function of the density of the county. Currently, given a population of ten thousand agents, the simulation creates densities of 44 agents per square km, 10 agents per square km, and 1 agent per square km. Next, the agents are instantiated. After all agents are created, undirected links are created between pairs of agents. After all links are created, all connected components are assigned to another agent that represents the location of their interaction. For example, if there are 5 agents connected together by school-type links, they will all be assigned to a single schoolhouse agent. The same is done for all other types of links. The final step is to take all agents not assigned to a home (those that live alone) and assign them to one.

Once all agents are assigned to a location agent, the location agents are then placed within the environment. This can be done in one of two ways. The first is to randomly place all location agents about the environment. The other way is to place all home agents on the right half of the environment and all non-home agents on the left side of the environment. This allows us to simulate counties that have segregated land use or integrated land use. Once that locations are placed, all links are removed to reduce the memory footprint of the simulation and a random selection of a user specified number of agents are exposed to the disease.

The initialization process can be time consuming when importing the inferred contact network. Therefore, once a population is created via the inferred network, the simulation can be reinitialized by resetting all parameter values, setting all agent health statuses to healthy, and resetting all agent locations to their home. This dramatically reduces the time required to run a design of experiment.

A time step is twelve hours in the model. Assuming no NPIs are activated, the agents spend odd time steps at home and even time steps at one of their non-home locations (if they have at least one of them). Currently, the model does not differentiate days of the week or times of year. During each time step individuals that are sick potentially spread the disease (additional details can be found in the disease propagation section). There are a number of NPIs that may be employed during a run to try and slow the spread of the disease. These include: social distancing, testing, contact tracing, closing schools, closing work sites, closing pubic venues, and isolating individuals.

Social distancing can be turned on or off at any point during a run. Once on agents have a user defined probability of staying at the home location rather than going to their non-home location during even time steps. This probability can be changed during runtime. Each location agent has a variable that designates it open or closed. If that variable is set to closed, all agents that wish to go to that location will remain in their current location rather than move to that location agent. The open variable of locations can be changed at any time during a run. A given run ends at a specified time step or when all individuals are in a state of healthy, recovered, or deceased.

#### Verification and Validation

Balci and Osman define verification as “substantiating that the model is transformed from one form into another, as intended, with sufficient accuracy” [23]. In the context of the ABM and COVID-19, the original form of the model is an extension of the canonical SEIR model where the assumptions of continuity are relaxed in favor of discrete agents and the concepts of behavioral changes induced by NPIs are incorporated. The final form of the model is a NetLogo representation of that logic. Therefore, in the absence of NPIs we would expect the ABM to re-produce the standard SEIR curves that are dominated by the decline in the Susceptible population and the increase in the Recovered and Deceased populations. This familiar set of curves is indeed reproduced by the ABM as illustrated in Fig. 2. Note the dark lines in the figure represent the mean of 30 simulation replications and the shaded areas represent one standard deviation above and below that mean.

**Fig 2.**
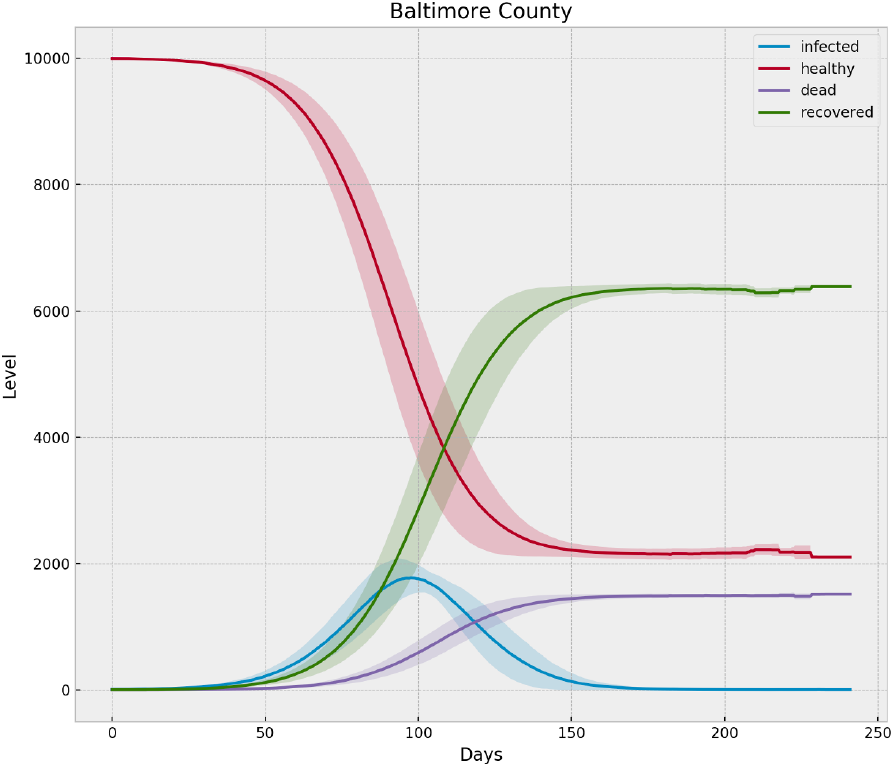
ABM Output with No NPIs for Baltimore County, MD

Validation is defined as “substantiating that the model, within its domain of applicability, behaves with satisfactory accuracy consistent with the [modeling and simulation] objectives” [23]. Balci and Osman note that accuracy assessment is validation when compared to the corresponding system behavior. For the run generating Fig. 3 we incorporate the NPIs of social distancing, testing, and tracing during the model run and note that the peak is lower than the baseline no-NPI run and the curve is rougher due to the more heterogeneous nature of social contact, which is what social distancing is designed to induce.

**Fig 3.**
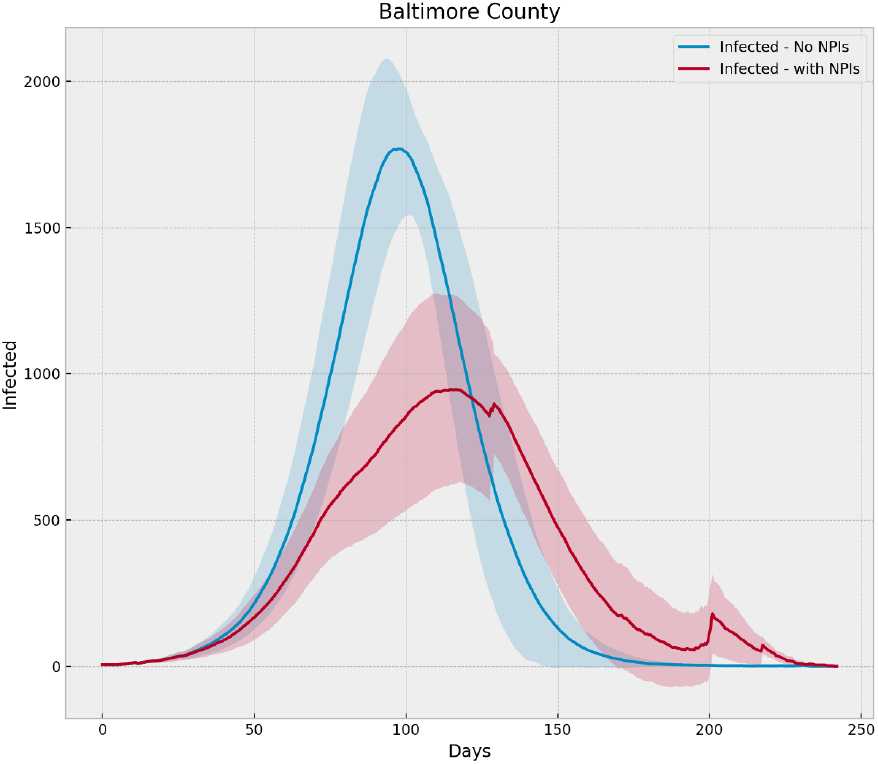
Comparison of ABM Output with and without NPIs for Baltimore County, MD

Proceeding in this manner we constructed a design of experiment to validate the other components of the model – such as testing and contact tracing – behave with satisfactory accuracy consistent with our modeling objective of assessing the risks associated with lifting NPIs. The heatmap shown in Fig. 4 shows the interaction of different levels of testing and contact tracing for a representative dense county with two-steps-away contact tracing. As expected the two tactics interact to progressively reduce the peak number of infections and deaths. Last, due to the way NetLogo currently ingests the graphical structure, we noted that runs performed with the same pseudo-random number seed are not exactly the same, i.e., reproducibility is not guaranteed. As a result, we base all our results on mean behavior rather than a specific replication of the simulation.

**Fig 4.**
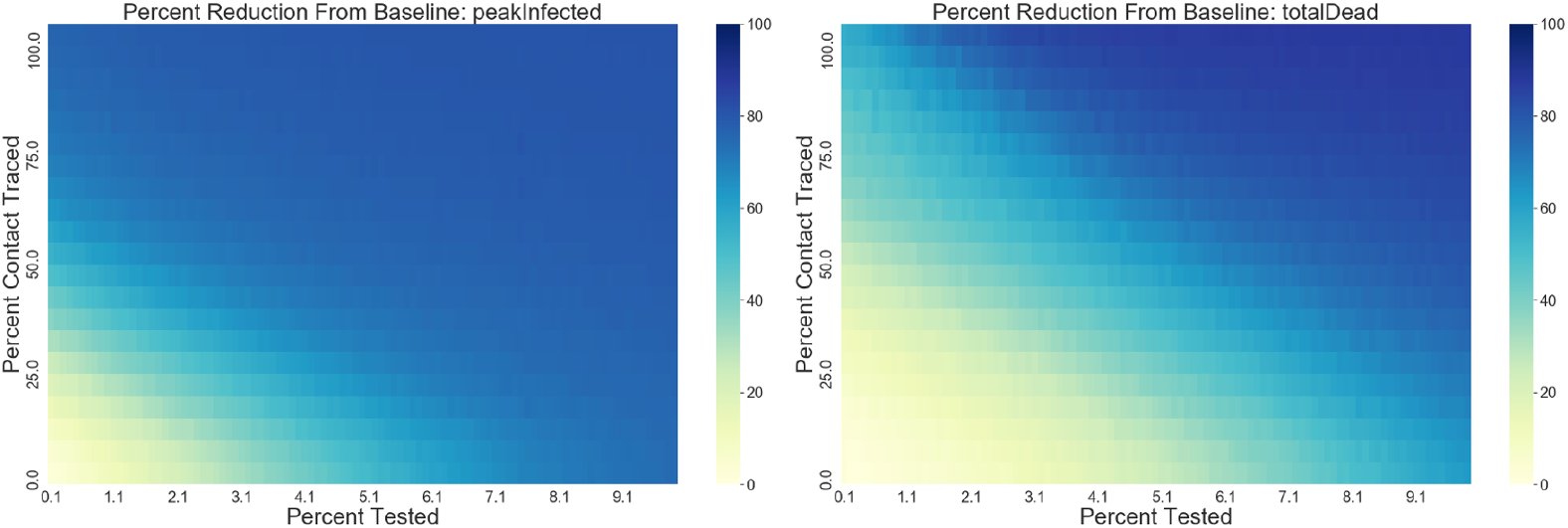
Heat Map of Test and Trace Design Points

Finally, we compared results from the simulation with the actual case counts reported in Maryland. This is a good exercise for ensuring model results are realistic, but it should be noted that precise statistical matches are *not* expected. There is uncertainty due to testing, the timing of NPIs, the actual adherence to NPIs, and relative scale of our 10,000 person simulations and the actual populations of a given county. Nevertheless the results shown in Fig. 5 and 6 indicate that our model results reasonably represent the counties they are intended to model.

**Fig 5.**
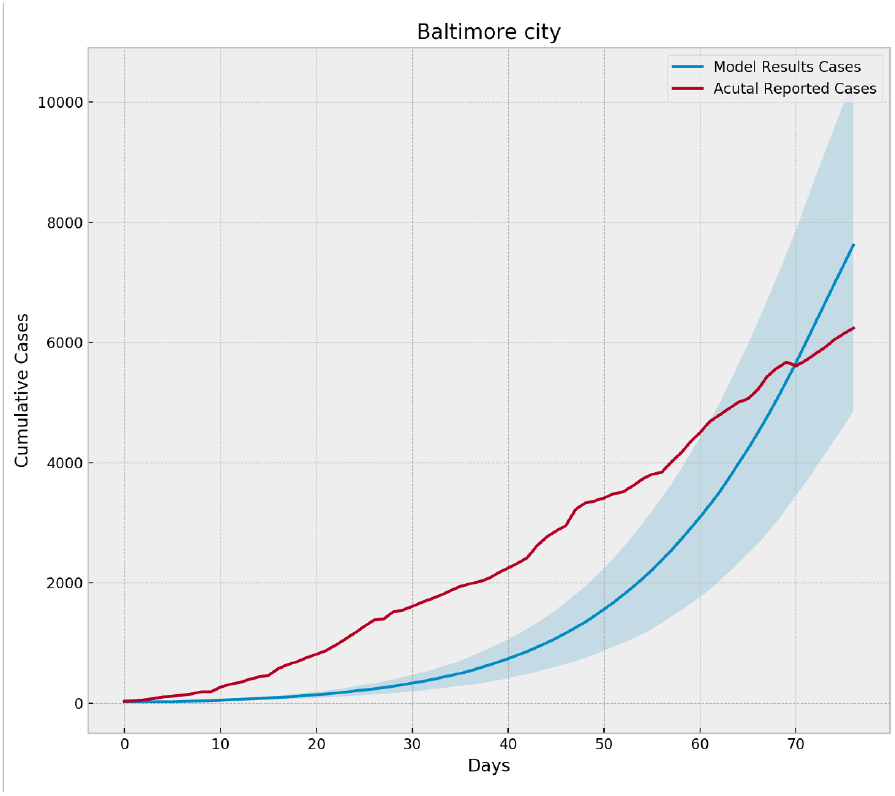
Comparison of Simulation Output to Actual Cases Reported in Baltimore City, MD

**Fig 6.**
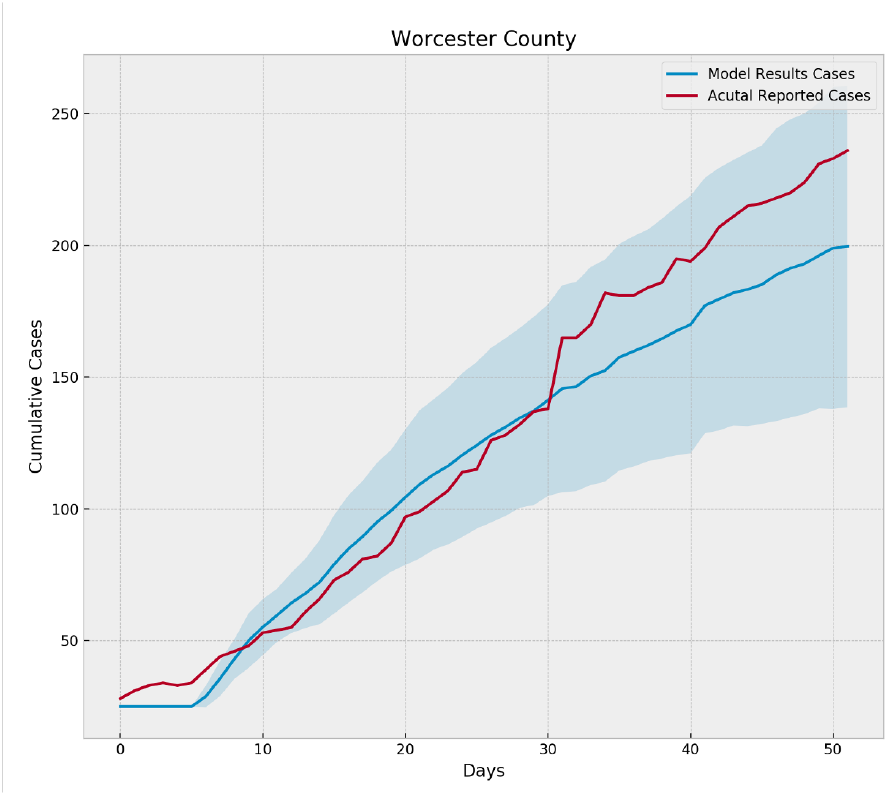
Comparison of Simulation Output to Actual Cases Reported in Worcester County, MD

#### Model Parameters and Sensitivity

As currently implemented the model contains a number of parameters. Most of the parameters are used to define the population (size) and its structure (spatial extent and mixing of home and non-home locations, size of the hospital, etc.). There are also a number of other parameters associated with the use of NPIs (when to turn them on or off). Finally, there are a number of parameters directly associated with the spread of the disease: transmission probability, mitigation probability, compliance with social distancing orders, compliance with isolation orders, and structure and extent of testing.

In our validation exercise the simulation demonstrated sensitivities one would expect to find in a model of this type. Disease spread was highly correlated to transmission probability and mitigation probability. Furthermore, testing made a significant difference when it was coupled with a population that complied with isolation orders. No unexpected sensitivities were uncovered, but it is important to note that SARS-Cov-2 is a novel virus and studies are producing new insights on a near daily basis. Parameterizing the model should thus remain an evolving exercise.

### Experiment Design

To illustrate the utility of our modeling approach, we chose to model the 24 counties of Maryland, including the independent city Baltimore. The experiment is designed to answer the question: *what is the impact of a given percent of the population being tested and a given level of participation in contact tracing if NPIs are lifted 70 days after the onset of the pandemic?* This is approximately the time frame that Maryland followed when lifting NPIs in reality.

Across the analytical model and the ABM, a collection of non-pharmaceutical interventions are implemented. The analytical model employs two distinct types of social distancing accomplished by removing vertices from the network and re-computing the degree distribution. The social distancing types are

- uniform social distancing - a given percentage of vertices are removed at random
- targeted social distancing - vertices that meet or exceed a given degree are removed

The targeted approach is equivalent to finding super-spreaders and requiring them to isolate. The uniform social distancing corresponds to the type of social distancing employed in the ABM. However, note that changing the degree distribution is essentially what venue closings and self-isolation requirements also accomplish in the statistical sense.

The NPIs included in the agent-based model are social distancing, isolation of infected individuals and individuals waiting for test results, and the closing of schools, workplaces, or public venues (e.g., grocery stores, shops, parks). In the ABM, all agents have a home location and spend at least half their time at that location (i.e., nights). Most agents also have a non-home location such as a workplace, a school, or public venue. Agents with non-home locations spend daytime hours at these locations. When social distancing is turned on, agents perform a random draw (compared against a user definable threshold) before leaving their home location. If the random number does not exceed the threshold, the agent stays home. This allows the user to vary the likely compliance to social distancing orders and to allow for agents to occasionally leave their home for necessities such as food and non-COVID healthcare. This is equivalent, in the limit, to uniform social distancing in the analytical model.

To implement isolation of infected individuals, those who test positive for COVID-19 in the ABM and those waiting for their test results (assumed to take three days in the simulation), are placed in isolation. When an agent is in isolation, they either stay at their home location or the hospital. These individuals will remain in isolation for two weeks when tested positive or three days when waiting for test results that come back negative. There is a user defined probability that an individual will comply with the isolation order.

Finally, homes, schools, workplaces, and public venues all have a specific location in the ABM and agents move from one of these locations to another depending upon time of day and their inferred contact network and demographic characteristics. With the exception of homes, these locations have a variable that allows them to be closed or opened. When closed agents will not go to them when they attempt to leave home. If the agent has no locations outside their home that are open, they will stay home. Any combination of venues can be closed, allowing for partial or phased re-openings to be simulated.

Maryland includes a mix of rural and urban counties. Baltimore and the suburbs of Washington, D.C. are the most populated areas with over 3 Million people, while Kent County has only around 19,434 inhabitants [8]. Table 3 lists the population, area, and density of each county according to the U.S. Census estimate in 2018.

**Table 3.** Population and Area of Maryland Counties.

| County | Population | Area (sq-km) | Density |
| --- | --- | --- | --- |
| Montgomery | 1048478 | 1368.31 | 766.26 |
| Prince George's | 909619 | 1306.65 | 696.15 |
| Baltimore | 827859 | 1646.66 | 502.75 |
| Baltimore city | 602443 | 219.23 | 2747.97 |
| Anne Arundel | 575523 | 1124.78 | 511.67 |
| Howard | 322621 | 685.24 | 470.81 |
| Frederick | 254943 | 1809.19 | 140.92 |
| Harford | 253882 | 1202.21 | 211.18 |
| Carroll | 168267 | 1228.06 | 137.02 |
| Charles | 161476 | 1234.90 | 130.76 |
| Washington | 150656 | 1269.91 | 118.64 |
| St. Mary's | 112720 | 970.63 | 116.13 |
| Wicomico | 103044 | 1009.31 | 102.09 |
| Cecil | 102644 | 960.84 | 106.83 |
| Calvert | 92065 | 577.42 | 159.44 |
| Allegany | 70941 | 1167.73 | 60.75 |
| Worcester | 51960 | 1265.22 | 41.07 |
| Queen Anne's | 50148 | 1006.06 | 49.85 |
| Talbot | 37074 | 724.2 | 51.19 |
| Caroline | 33306 | 861.95 | 38.64 |
| Dorchester | 31960 | 1514.71 | 21.10 |
| Garrett | 29133 | 1779.60 | 16.37 |
| Somerset | 25606 | 873.70 | 29.31 |
| Kent | 19434 | 760.43 | 25.56 |

Using the U.S. Census ACS data and the algorithm described earlier, we constructed the 24 contact structure graphs to represent the pre-NPIs state for initializing each simulation. The parameters used in constructing the graphs are shown in Table 4. These graphs were ingested into the ABM as the initial contact structure. The degree distributions of each county graph are characterized by the means and standard deviations listed in Table 5.

**Table 4.**
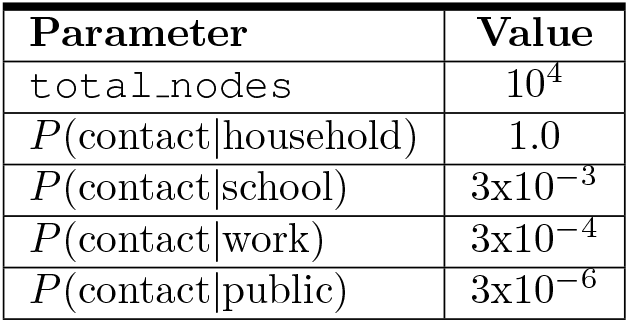
Parameters for Social Contact Structure.

**Table 5.** Degree Distributions of Social Contact Structures by County.

| County | Mean | St. Dev |
| --- | --- | --- |
| Allegany County | 2.49 | 2.02 |
| Anne Arundel County | 2.72 | 2.09 |
| Baltimore County | 2.67 | 2.10 |
| Calvert County | 2.91 | 2.41 |
| Caroline County | 2.75 | 2.53 |
| Carroll County | 2.72 | 2.10 |
| Cecil County | 2.74 | 2.24 |
| Charles County | 2.95 | 2.60 |
| Dorchester County | 2.40 | 2.13 |
| Frederick County | 2.83 | 2.14 |
| Garrett County | 2.23 | 1.85 |
| Harford County | 2.67 | 2.05 |
| Howard County | 2.97 | 2.21 |
| Kent County | 2.44 | 2.23 |
| Montgomery County | 2.80 | 2.11 |
| Prince Georges County | 2.97 | 2.90 |
| Queen Annes County | 2.58 | 2.05 |
| St. Marys County | 2.96 | 2.28 |
| Somerset County | 3.10 | 4.48 |
| Talbot County | 2.05 | 1.75 |
| Washington County | 2.58 | 2.15 |
| Wicomico County | 3.14 | 2.82 |
| Worcester County | 2.10 | 1.86 |
| Baltimore city | 2.71 | 2.50 |

The full experiment consisted of three sets of simulation runs. The first was a baseline run of 30 replications for each county with no NPIs or interventions. This scenario essentially represents the baseline course of the pandemic if no action were taken to slow the spread of disease. The second scenario instituted multiple NPIs 10 days after the start of each run and then partially lifted those NPIs 70 days after the start of each run. The NPIs were the closing of school and workplace venues, but not public venues. Social distancing was also enforced starting on day 10 at 95% and then reduced to 50% after 70 days. When the NPIs were lifted a regime of testing and contact tracing was instituted. For this particular set experiment we employed one-step contact tracing. That is, agents who came in direct contact with an infectious agent and opted-in to the program were traced. But agents who came in contact with an agent who was traced were not in turn traced. In this scenario only symptomatic people were tested and contact tracing commenced for those cases that tested positive and had opted-in to the tracing program. The symptomatic group includes only those individuals in the Mild, Severe or Critical states. Five levels of testing were used and five levels of agreeing to isolate after contact tracing (`optInRate`) were used for a full design of experiment consisting of 25 settings and 30 replications per county. The third scenario uses the same NPIs and timing of NPIs as the second scenario, but employs a random testing regime. This includes agents that are in the Susceptible, Exposed, Mild, or Recovered states. The same five levels of `optInRate` were used, but the percent tested was varied across a set of higher testing levels. In our exploratory experimentation we found that higher percentages of random testing are required to achieve similar impact because the same quantity of tests uncovers fewer true-positives. Tables 6, 7, and 8 outline the parameter settings for each of the three scenarios. Note that by setting social distancing to a high percentage but leaving venues open, we simulate minimal interaction that occurs at essential businesses such as grocery stores.

**Table 6.**
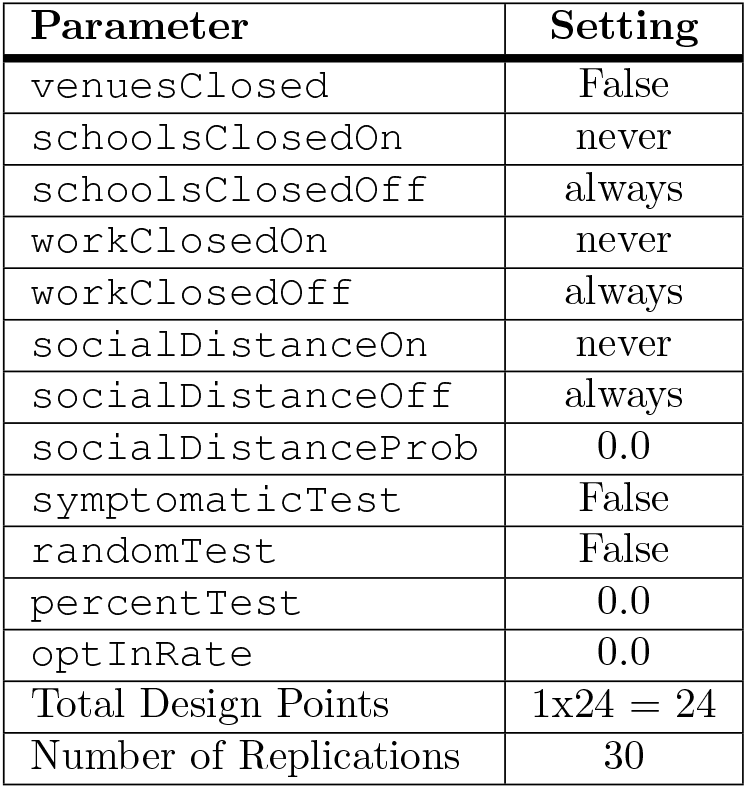
Parameter settings for the baseline scenario.

| Parameter | Setting |
| --- | --- |
| `venuesClosed` | False |
| `schoolsClosedOn` | never |
| `schoolsClosedOff` | always |
| `workClosedOn` | never |
| `workClosedOff` | always |
| `socialDistanceOn` | never |
| `socialDistanceOff` | always |
| `socialDistanceProb` | 0.0 |
| `symptomaticTest` | False |
| `randomTest` | False |
| `percentTest` | 0.0 |
| `optInRate` | 0.0 |
| Total Design Points | $1 \times 24 = 24$ |
| Number of Replications | 30 |

**Table 7.** Parameter settings for Symptomatic Testing scenario.

| Parameter | Setting |
| --- | --- |
| `venuesClosed` | False |
| `schoolsClosedOn` | step 20 (day 10) |
| `schoolsClosedOff` | step 140 (day 70) |
| `workClosedOn` | step 20 (day 10) |
| `workClosedOff` | step 140 (day 70) |
| `socialDistanceOn` | step 20 (day 10) |
| `socialDistanceOff` | never |
| `socialDistanceProb` | [0 to step 20, 0.95 to step 140, 0.5 to end] |
| `symptomaticTest` | True |
| `randomTest` | False |
| `percentTest` | [0.002, 0.004, 0.006, 0.008, 0.01] |
| `optInRate` | [0.0, 0.25, 0.5, 0.75, 1.0] |
| Total Design Points | $5 \times 5 \times 24 = 600$ |
| Number of Replications | 30 |

**Table 8.**
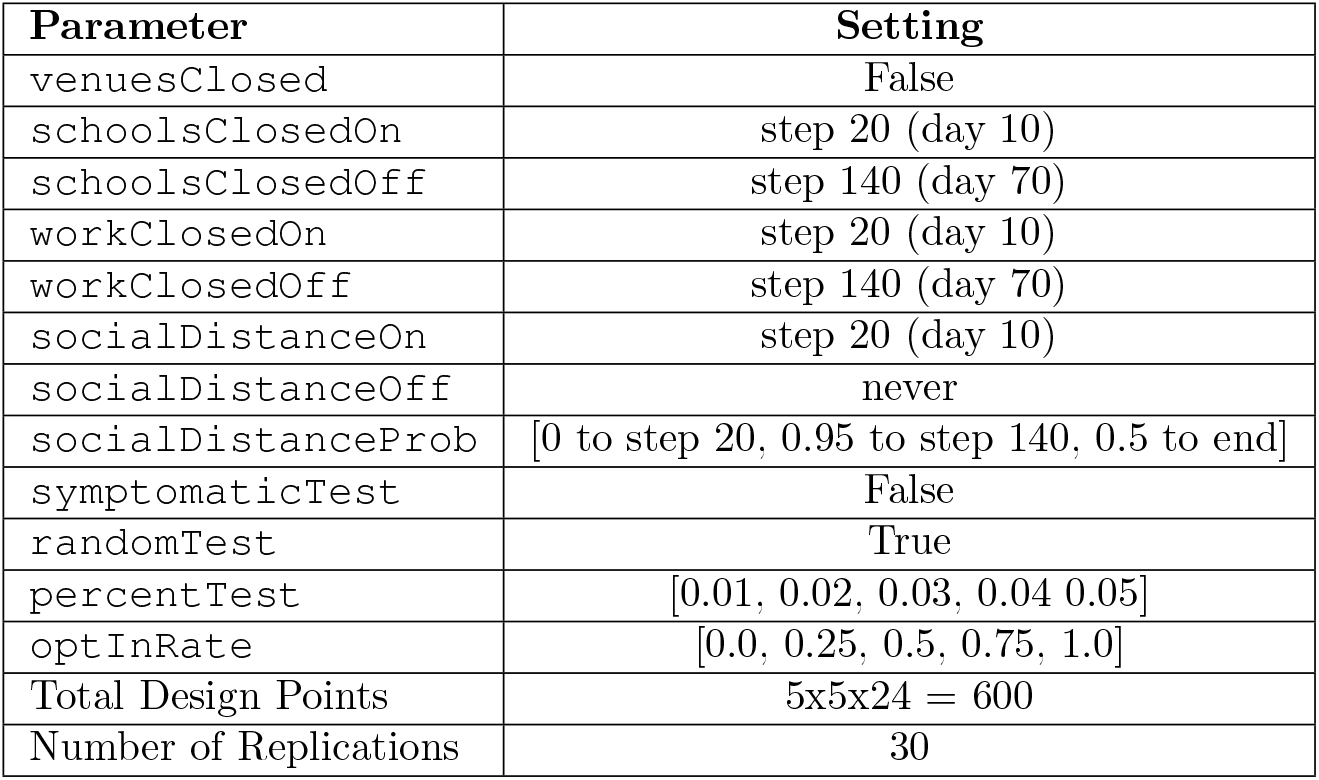
Parameter settings for Random Testing scenario.

| Parameter | Setting |
| --- | --- |
| venuesClosed | False |
| schoolsClosedOn | step 20 (day 10) |
| schoolsClosedOff | step 140 (day 70) |
| workClosedOn | step 20 (day 10) |
| workClosedOff | step 140 (day 70) |
| socialDistanceOn | step 20 (day 10) |
| socialDistanceOff | never |
| socialDistanceProb | [0 to step 20, 0.95 to step 140, 0.5 to end] |
| symptomaticTest | False |
| randomTest | True |
| percentTest | [0.01, 0.02, 0.03, 0.04 0.05] |
| optInRate | [0.0, 0.25, 0.5, 0.75, 1.0] |
| Total Design Points | 5x5x24 = 600 |
| Number of Replications | 30 |

## Results

### Agent-based Model Results

In this section we review and compare the results of the different scenarios. It is important to note that these results do *not* represent a forecast of case counts or death rates. The purpose of this modeling approach is to provide insight into the relative impact that different NPI, testing, and tracing strategies will have when applied to areas of a particular state that have considerably different social contact structures.

Fig. 7 shows the impact of population density and social contact structure. Each box-whisker plot represents the distribution of the maximum number of infections across the 30 replications of the baseline scenario for a given county. Recall that the disease parameters are the same for each county and the populations are normalized to 10^4^. The two variables that change from county-to-county are only the social contact structure driven by the U.S. Census data, and the population density controlled by the size of the environment the agents have to move around in. The key insight is that any strategy the state of Maryland adopts will need to treat 7 of the 24 counties differently. Referring to Table 3, it is interesting to note that Harford County has roughly 75% fewer people than Montgomery County and Montgomery County is roughly 70% less dense than Baltimore City. Yet all three of these locations have an average maximum infected that is approximately four times larger than 17 of the counties in rest of the state.

**Fig 7.**
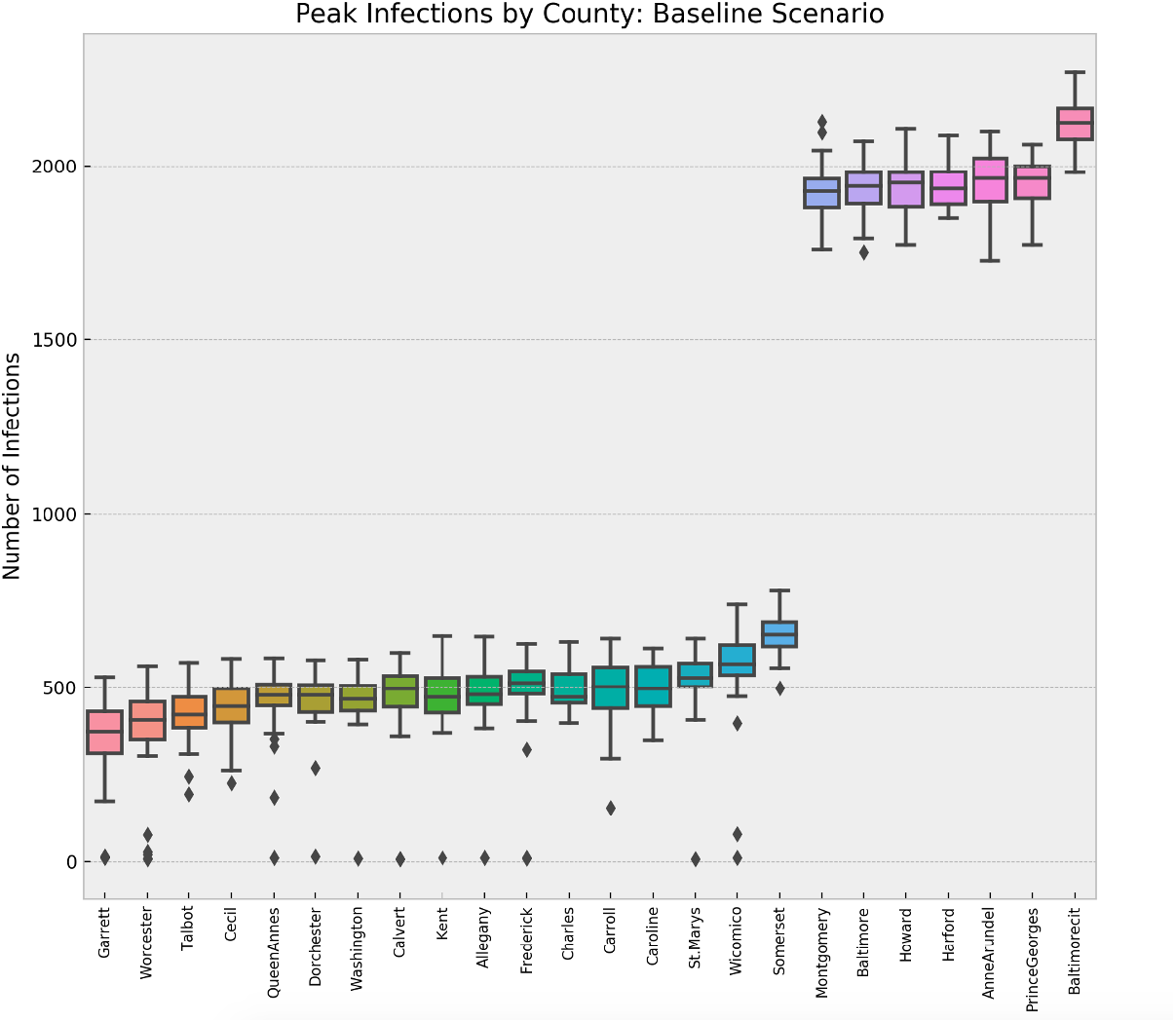
Mean Peak Infections across Replications by County

Next we analyzed the impact of a 50-day NPI strategy followed by a regime of symptomatic testing and contact tracing. From a broad perspective we can see in Fig. 8 and Fig. 9 that the NPIs and testing and tracing combine to reduce the total cumulative cases considerably. Here we can see that if each county has the ability to test 1% of its population daily then the overall number of cases can be drastically reduced. Also note that, for this level of testing and contact tracing, the dynamics in Baltimore City separate from the other counties, most likely due to its extremely high density. There is still a difference from one county to the next due to the differences in density and contact structure. We selected three of the counties to look at in greater detail that are notionally representative of high, medium, and low density locations.

**Fig 8.**
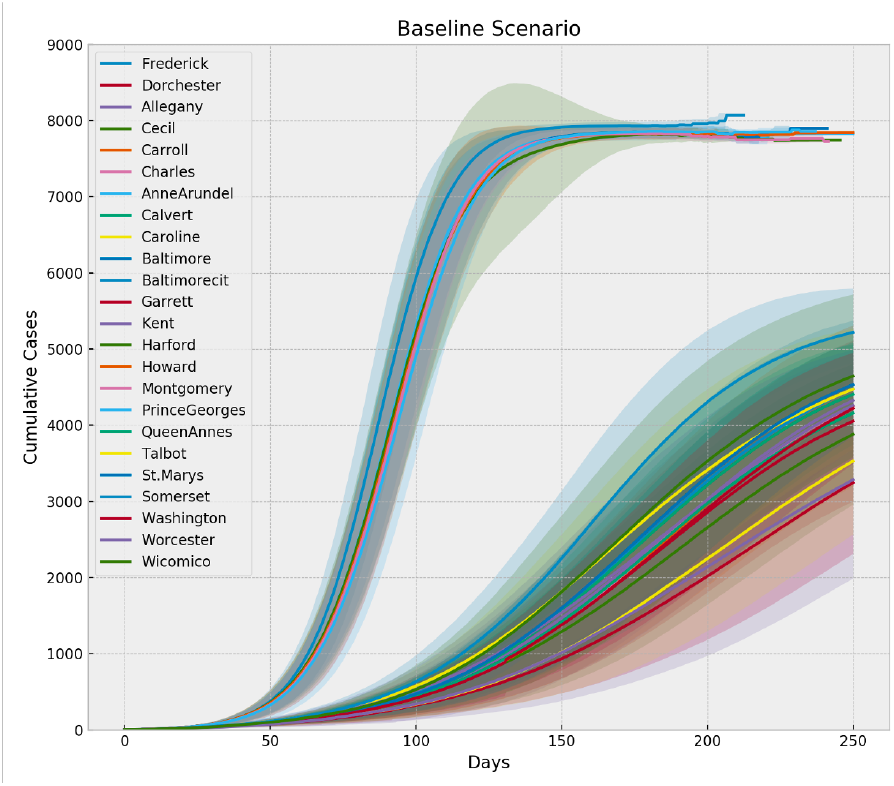
Cumulative Cases for Baseline Scenario

**Fig 9.**
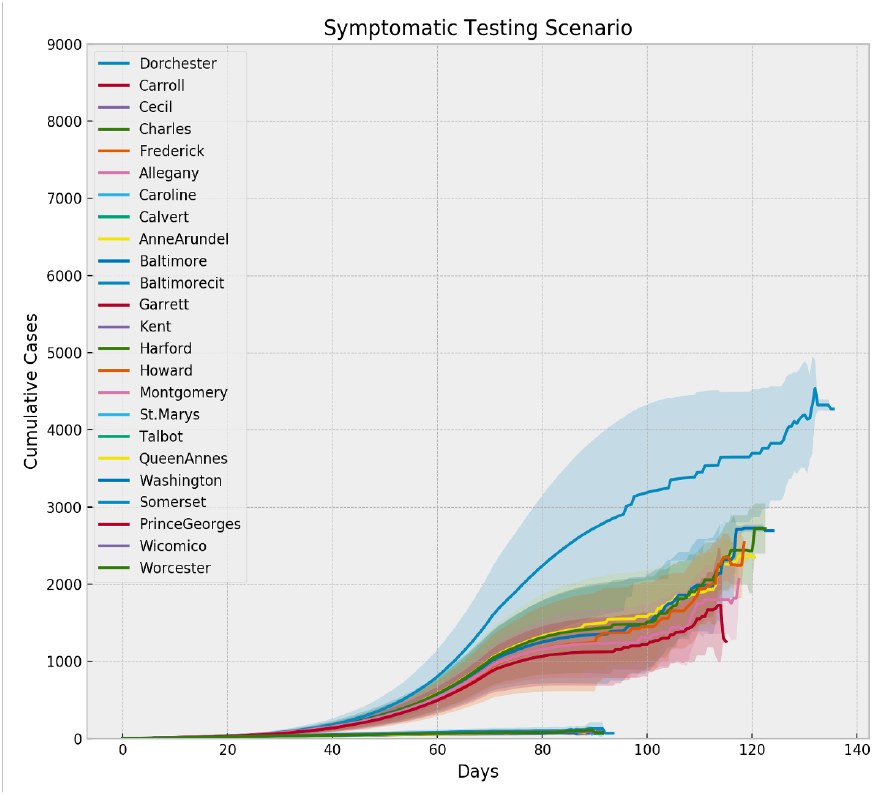
Cumulative Cases for Symptomatic Testing Scenario (0.01 test, 1.0 `optInRate`)

Baltimore City has the highest population density out of the 24 locations. Harford is seventh out of 24 in terms of density and Worcester is near the bottom. Fig. 10, 11, and 12 show the mean peak infections for each of the 25 design points of the symptomatic testing scenario, along with the baseline mean peak infections for each of these counties. The combined impact of the NPIs, testing, and tracing is evident, but the variation in the impact is greatly affected by the density and social contact structure of the county. It is less obvious from these plots whether different levels of testing or contact tracing have a significant effect on the mean peak infections. Indeed, using a two-sample Kolmogorov-Smirnov test with *α* = 0.05 we can only reject the null hypotheses that the mean peak infections are the same when testing the baseline against the COVID-19 testing level of 0.002 and comparing that level of testing with any other level of testing for Baltimore and Harford Counties. In Worcester County all levels of testing are distinguishable from the baseline, but not from each other.

**Fig 10.**
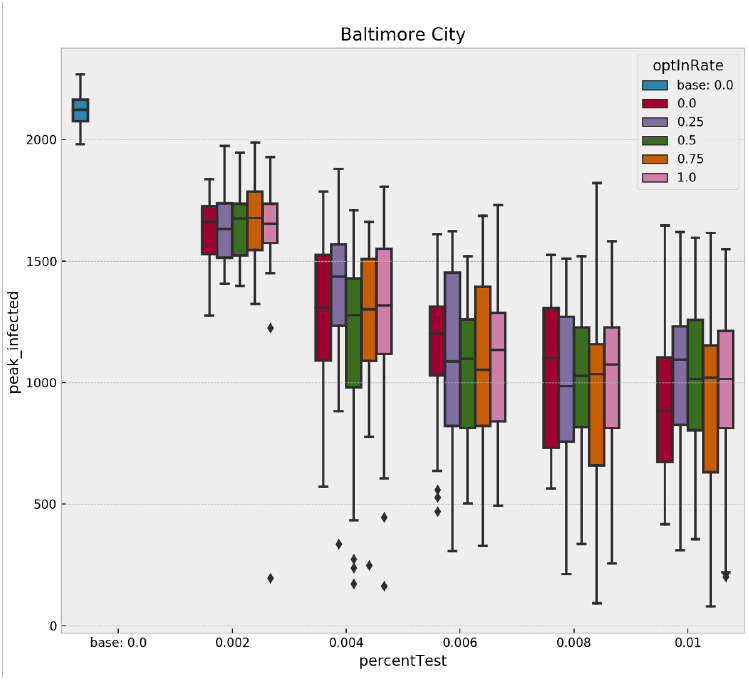
Baltimore City Scenario Comparison

**Fig 11.**
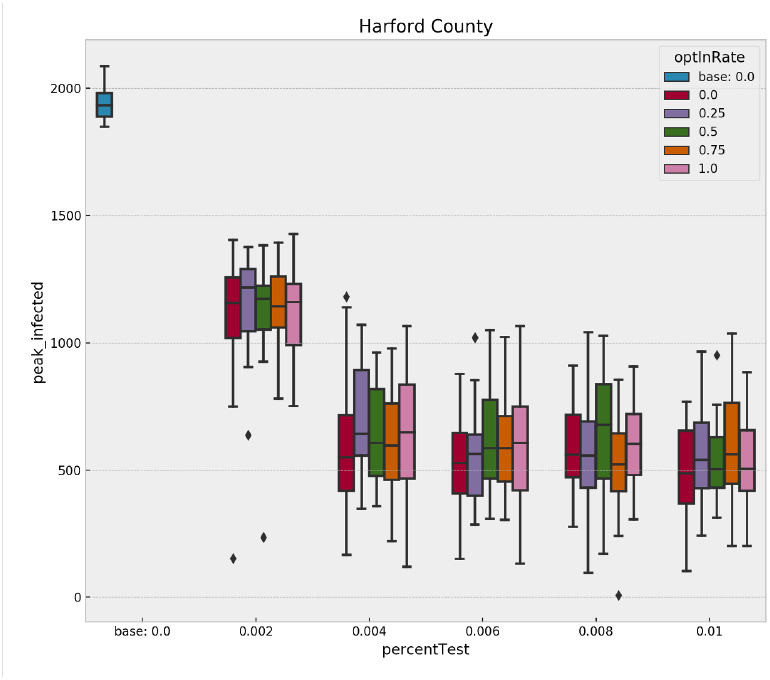
Harford County Scenario Comparison

**Fig 12.**
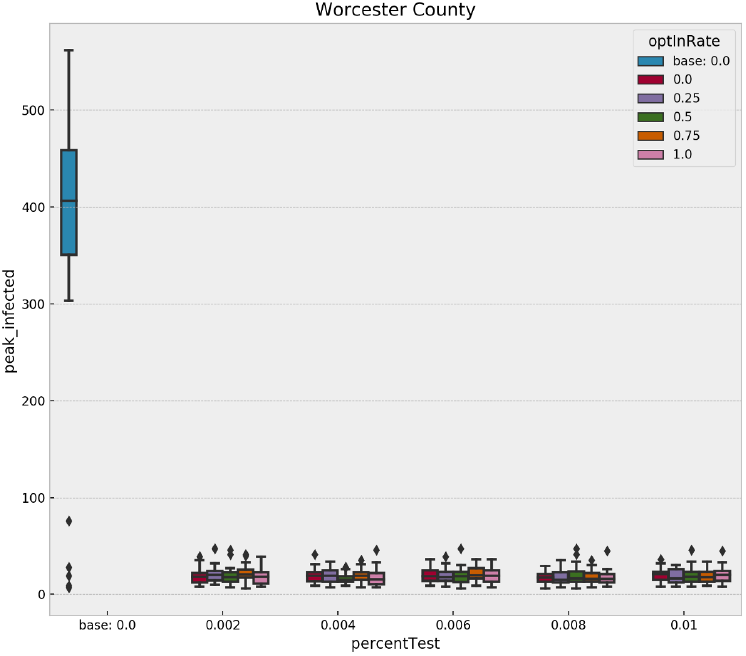
Worcester County Scenario Comparison

Interestingly, the story changes when we analyze deaths in these three counties within the 250 simulated days, as seen in Fig. 13, 14, and 15. The differences between certain levels of testing becomes more pronounced. Again, applying a two-sample Kolmogorov-Smirnov test to the different pair-wise combinations of testing levels we can now differentiate all pairs except the highest two levels in Baltimore and Harford County (*α* = 0.05). The levels remain indistinguishable in Worcester County. These results are important for decision-makers trying to allocate scarce resources, such as testing and contact tracing, across multiple regions of interest because the same results can be obtained in some places with fewer resources than in others.

**Fig 13.**
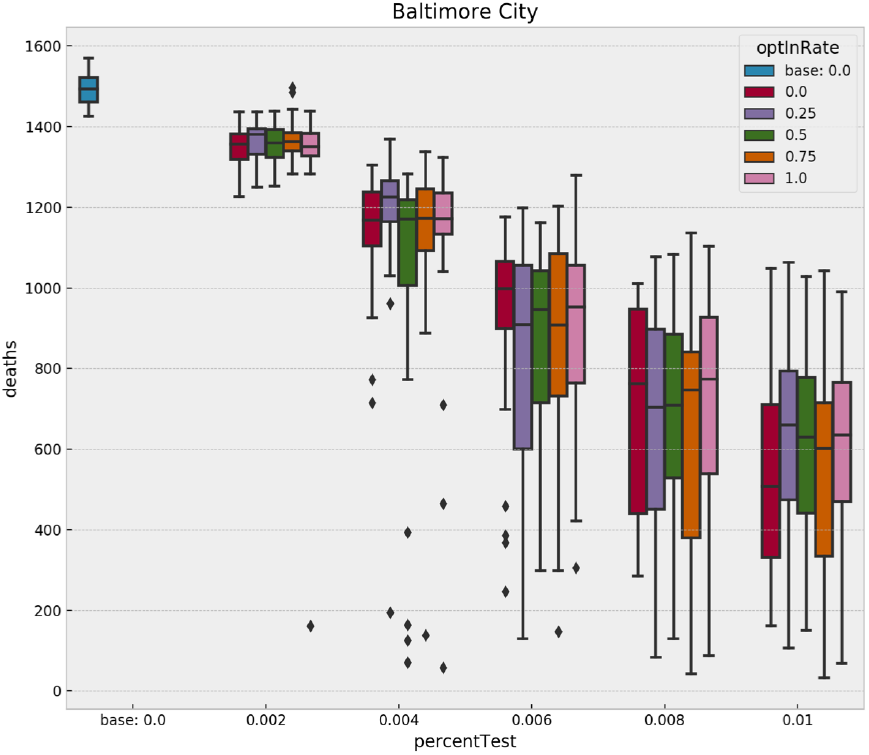
Impact of Symptomatic Testing and Tracing to Deaths in Baltimore

**Fig 14.**
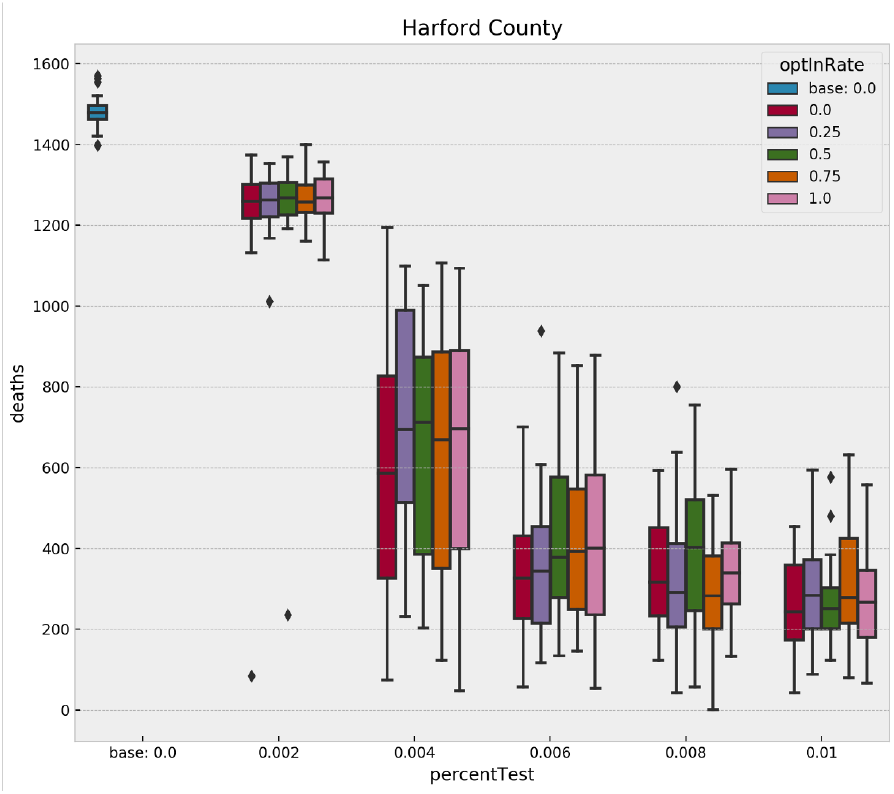
Impact of Symptomatic Testing and Tracing to Deaths in Harford County

**Fig 15.**
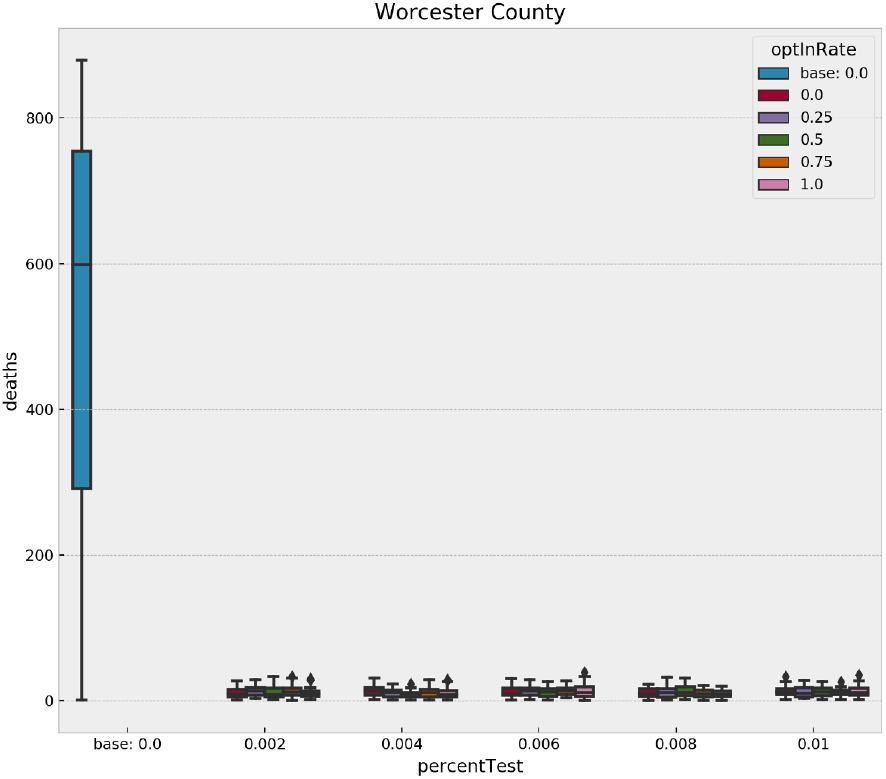
Impact of Symptomatic Testing and Tracing to Deaths in Worcester County

If we focus on any one county, we can analyze the interaction effects of different levels of testing and contact tracing, as well as if symptomatic or random testing produce different results. Recall that five of our design points between symptomatic testing and random testing overlap. Specifically, for all five levels of `optInRate`, the 1.0% testing level is included in both experiments. Holding the `optInRate` constant at 0.75, we can see in Fig. 16 that more than 5.0% random testing is required to achieve the same results at 1.0% symptomatic testing. This result may seem misleading at first because healthcare professionals agree that more testing and random testing of asymptomatic people is highly recommended. It is important to note that *all* symptomatic agents in the model are indeed infected with COVID-19. So 1.0% testing of symptomatic individuals is testing a large percentage of the infectious population. Conversely, the random testing regime is forced to distribute the tests across a mix of infectious and non-infectious people. Since the non-infectious population is larger when NPIs are employed, the diluted number of true-positive tests makes the random regime appear less effective at higher levels of testing. In actuality, this reinforces the message of increased random testing. We know that many COVID-19 cases often exhibit few symptoms even though the individuals are infectious. These individuals are less likely to submit for testing because they might not even know they are sick. Increased levels of random testing provides greater opportunity of finding and isolating those cases – as illustrated by the model results – but that also means a greater level of testing is required to actually find those who are infected. It is also important to note that random testing is required to estimate the prevalence of the disease.

**Fig 16.**
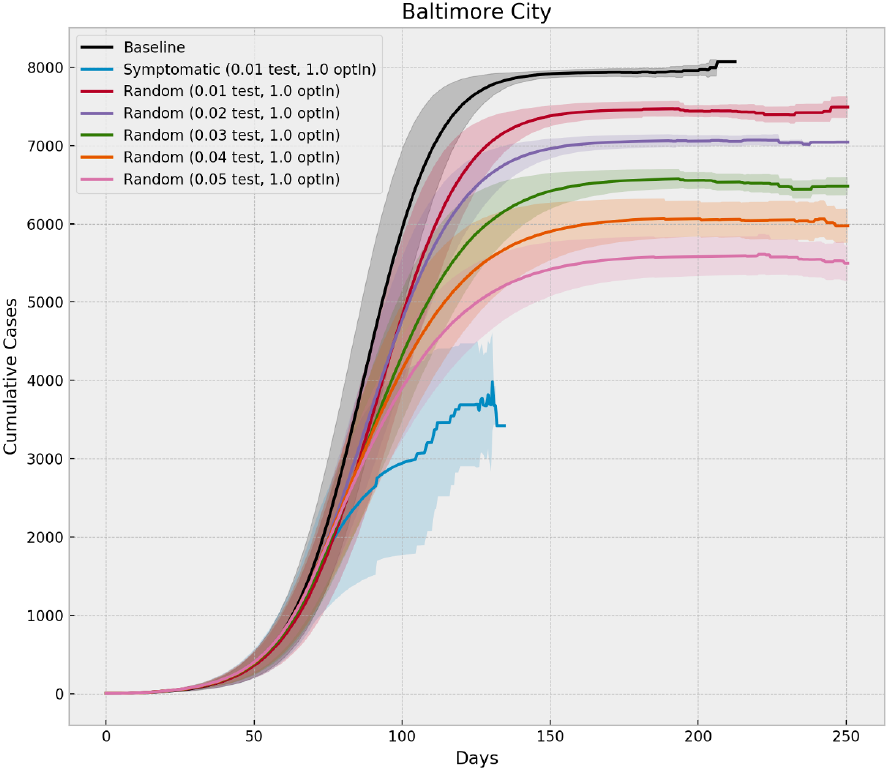
Cumulative Cases in Baltimore City under Different Testing Regimes

We can also analyze interaction effects through heatmaps that illustrate the two control variables across all 25 combinations. The mean peak infections in Baltimore for both symptomatic and random testing are shown in Figures 17 and 18. Not surprisingly, increased testing and increased participation in contact tracing (upper right corner) show a marked decrease in the peak number of infections.

**Fig 17.**
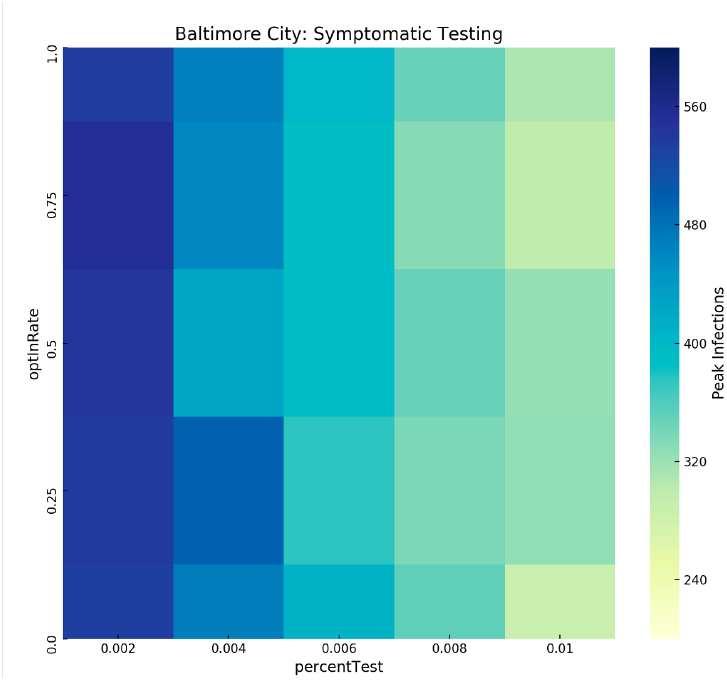
Baltimore Peak Infections by Design Point: Symptomatic Testing

**Fig 18.**
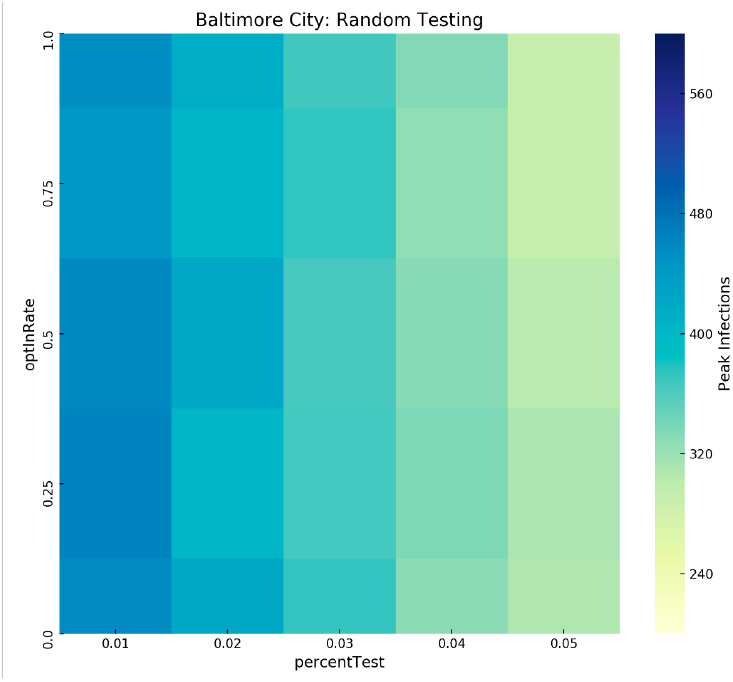
Baltimore Peak Infections by Design Point: Random Testing

This generally holds true for the other counties, but again the influence of contact structure and density can lead to a less monotonically decreasing relationship as illustrated in Fig. 19. In the county heatmaps we see that testing and contact tracing have less well defined results for rural counties. This is largely due to the structure of these communities and the fact that the number of infected individuals is small (per 10,000 people). As such the probability of random testing finding an infected individual is also small. This leads to fewer infected agents across all design points.

**Fig 19.**
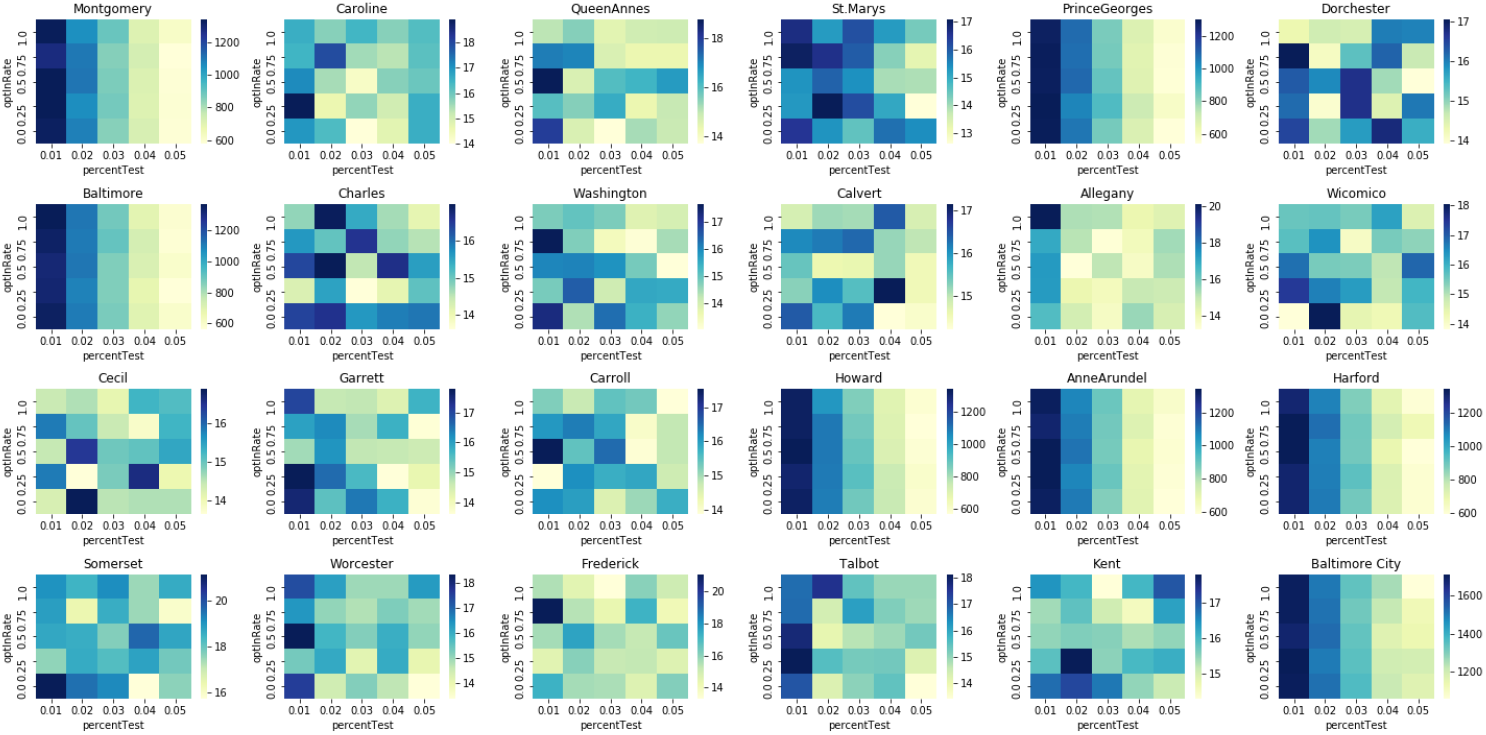
All Counties Random Testing Regime

For the designs of experiments presented here we saw little impact from contact tracing. We hypothesize that this is due to the simulations falling roughly into two buckets:

1. Too little testing to detect and quarantine enough individuals to have an impact. In this case contact tracing will have little effect because not enough cases are discovered.
2. Enough testing to bend the curve and stop the outbreak. In this case contact tracing is not required because there is already enough testing.

Further experiments could be conducted with a finer level of variation in testing to find the regime where contact tracing would have the most impact.

### Comparison with the Analytical Model

Previously we outlined a set of analytical models for estimating dynamic and steady-state disease spread in social contact networks using the theory of bond percolation. Comparisons of model results with ABM simulations are now examined for three counties in Maryland: Baltimore City, Somerset, and Worcester. Baseline results are initially compared with no social distancing, testing, or contact tracing. The network structure and degree distributions for each county are inferred from Census data and Results are scaled by the inferred population density of each county.

Figures 20, 21, and 22 compare the baseline number of infections over time as predicted by the ABM and analytical models. Analytical results are comparable to ABM (data) results for the three baseline cases examined. Both the maximum number of infections and the peak outbreak times are well predicted by the analytical model. There are small differences in the tail characteristics, but these differences are within the envelope of statistical uncertainty (*±* 1*σ*) based on the number of ABM simulation runs (30 replications).

**Fig 20.**
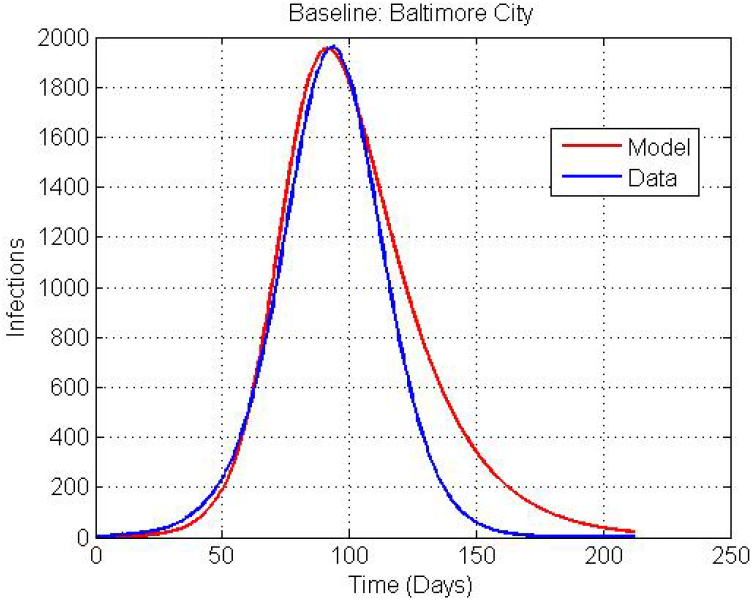
Analytical model comparison with baseline ABM simulations for Baltimore City, MD assuming no social distancing, testing, or contact tracing

**Fig 21.**
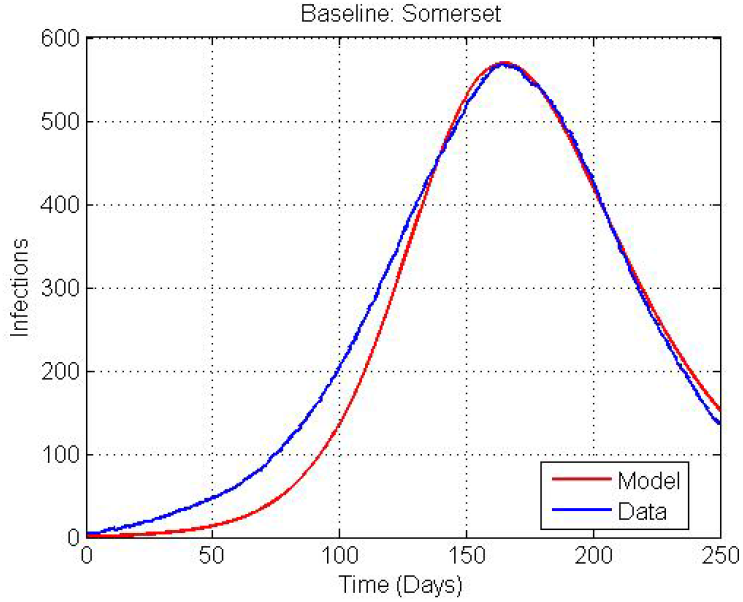
Analytical model comparison with baseline ABM simulations for Somerset County, MD assuming no social distancing, testing, or contact tracing

**Fig 22.**
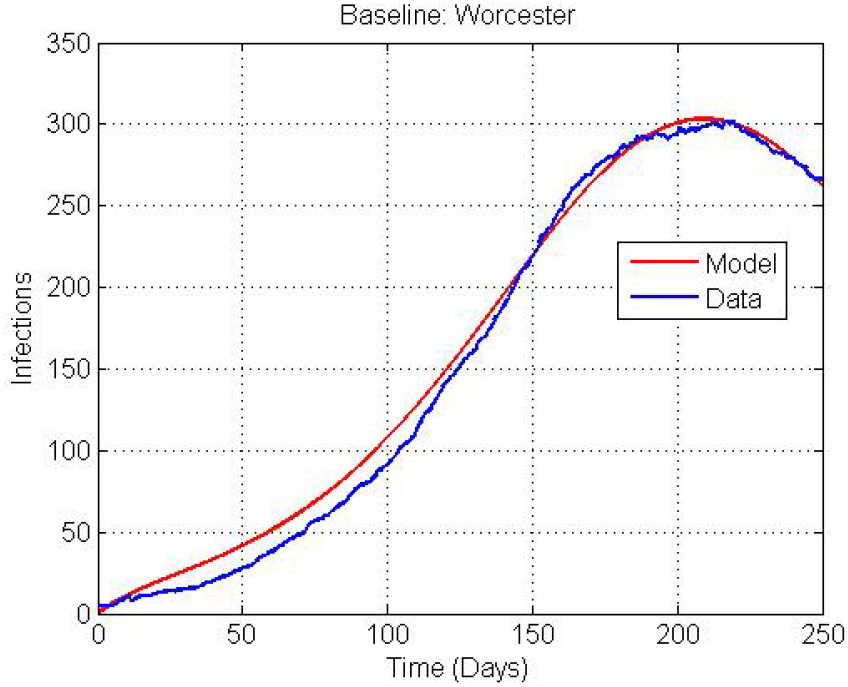
Analytical model comparison with baseline ABM simulations for Worcester County assuming no social distancing, testing, or contact tracing

Results are now compared when social distancing, testing, and contact tracing protocols are implemented. To briefly summarize, there is no social distancing up to day Between days 10-70, it is assumed that 95% of the population is social distanced. After day 70, 50% of the population is social distanced. Randomized testing and contact tracing are assumed starting at day 70. Fig. 23 is a comparison of ABM and analytical models for Baltimore City. Only 1% of individuals are randomly tested with a 50% opt-in rate for contact tracing. Analytical model results are comparable to the ABM model, including the peak number of infections and peak outbreak time.

**Fig 23.**
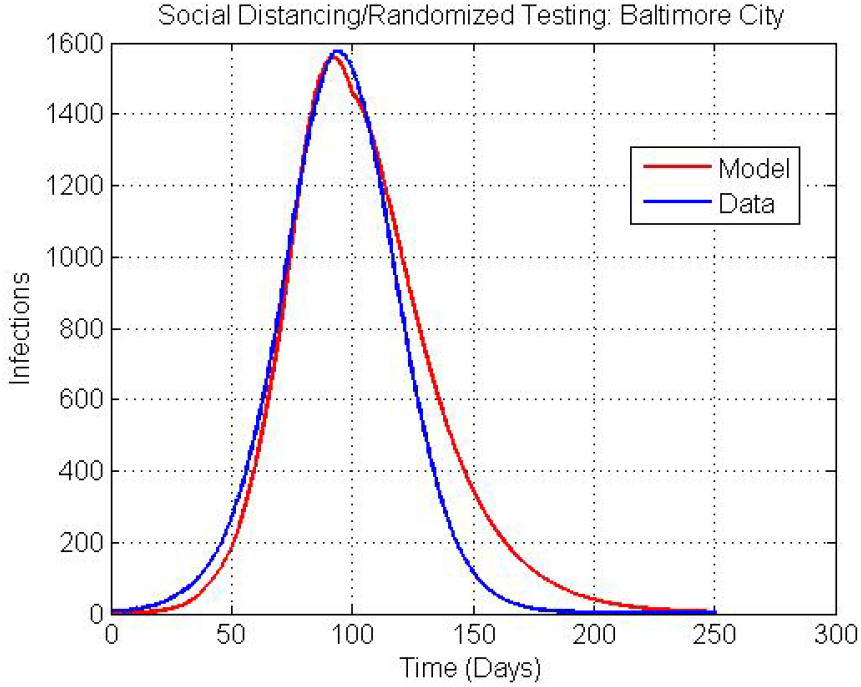
Analytical model comparison with ABM simulations for Baltimore City assuming social distancing and randomized testing and contact tracing

The results for Somerset and Worcester counties are shown in figures 24 and 25, respectively. For both counties, 4% of individuals are randomly tested with a 50% opt-in rate for contact tracing. Note that the ABM results are random in appearance because the number of infections is fairly small. However, the analytical model appears to approximately match the two peaks in the Somerset county infections predicted by the ABM model. Additionally, the analytical model approximately matches the Worcester county infections – the two peaks are in the correct location and the tails are within the statistical uncertainty of the ABM simulations.

**Fig 24.**
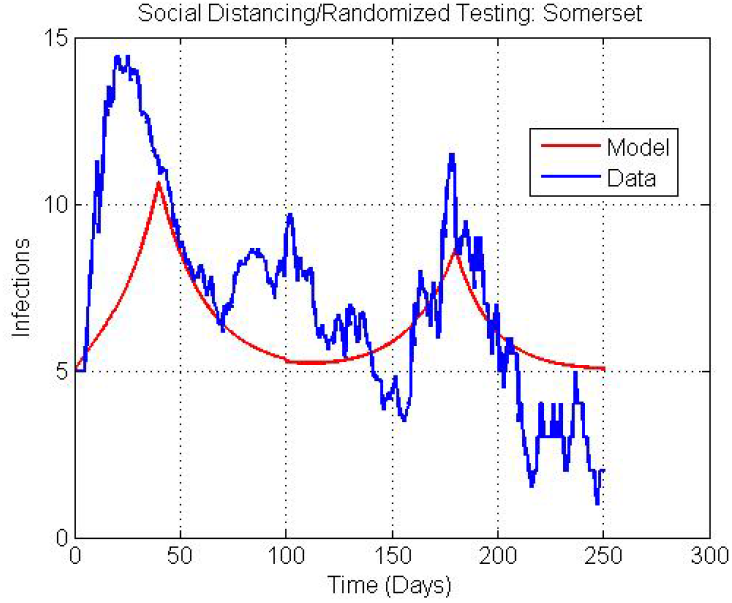
Analytical model comparison with ABM simulations for Somerset County assuming social distancing and randomized testing and contact tracing

**Fig 25.**
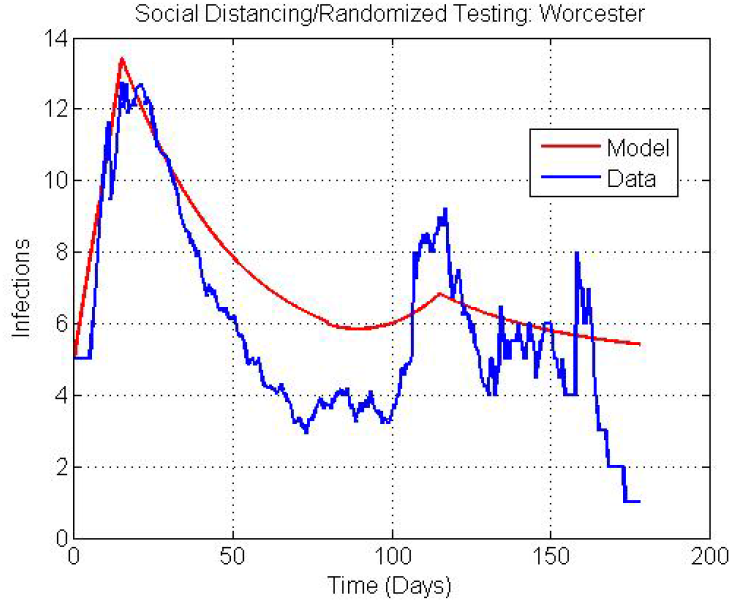
Analytical model comparison with ABM simulations for Worcester County assuming social distancing and randomized testing and contact tracing

Initial conclusions suggest that the analytical models can provide a complementary approach for the coarse-grain prediction of disease spread in social contact networks when rapid turnaround is required with low computational overhead. A more fine-grain analysis is afforded by the ABM model when detailed predictions are required.

## Discussion

In the present work, we illustrated an ensemble modeling approach (shown graphically in Fig. 26) for assessing alternate strategies of implementing and subsequently lifting non-pharmaceutical interventions in response to the COVID-19 pandemic. We underscored the previously-known result that social contact structure is a key factor in the size of an outbreak or pandemic and we illustrated how closed-form analytical models, network diffusion models, and agent-based models can be used in concert to provide insight to decision-makers in the face of uncertainty. We summarized our findings in a limited design of experiments that focused on the 24 counties and county-equivalents of the state of Maryland. We showed that the different counties of Maryland fall into at least two distinct categories in terms of risk of large outbreaks and illustrated how different levels of testing can be employed to the same effect if the social contact structure is taken into account. Finally, we showed that our analytical model produces a coarse-grain assessment that is in line with the ABM results. This approach is faster and less computationally expensive when rapid decision-making is necessary.

**Fig 26.**
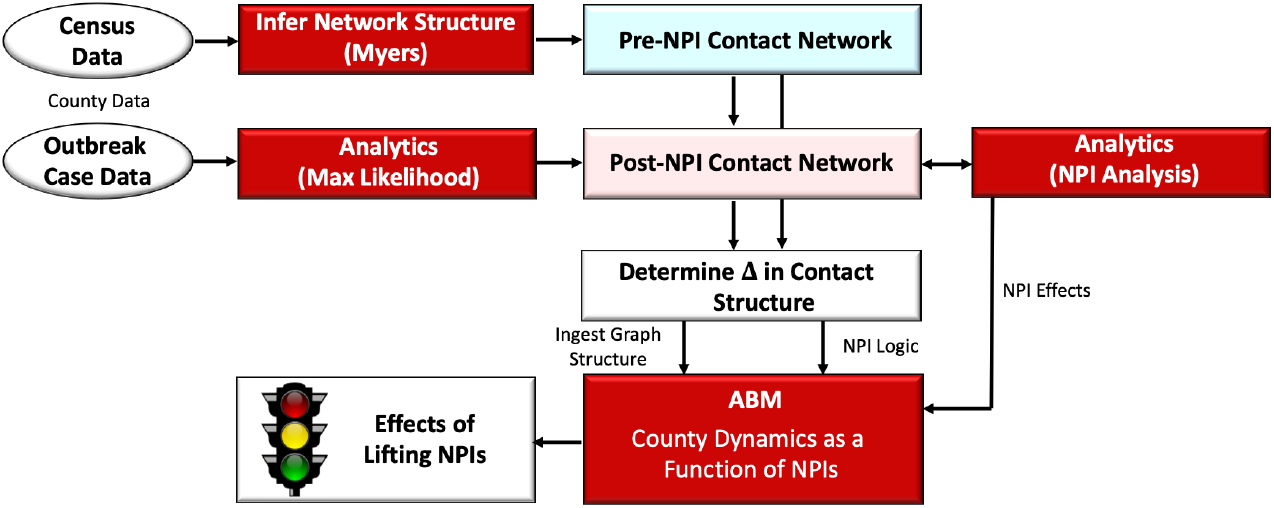
The ensemble approach work flow

It is important to note that no model or suite of models is a panacea. Ultimately decision-makers are forced to make a choice under uncertainty to protect both the health and the economic well-being of the citizenry. The approach outlined here is designed to provide insight into the marginal impact of one NPI and testing strategy versus another. This approach should be utilized by the decision-makers in conjunction with empirical analysis of the current state of their county or region of interest. The model parameters or logic should be constantly updated and the models re-run with new information as it comes available. That is, this modeling approach is designed for use in real-time alongside decision-makers at the time decisions are being formulated and implemented. To that end, the analysis presented here should be taken as notional rather than indicative of what might or might not happen in Maryland over the coming months.

During the writing of this report, unforeseen events extraneous to COVID-19 led to social unrest, protests, and riots in many major cities across the United States. Most of these locations were still operating under some level of restrictions to control the pandemic. Clearly, protests and riots bring people into close proximity and may ultimately prove to be super-spreading events. This sort of unpredictable event is not included in our model nor have we seen them included in the models we reviewed. This serves to underscore the difficulty and challenges of forecasting the progression of complex systems. Models and the insights they provide can help, but they are ultimately limited by assumptions. Decision-makers therefore require a combination of reliable data from their region of interest, rigorously designed models that make as few simplifying assumptions as possible, and ultimately the fortitude to make a decision in the face of uncertainty.

## Data Availability

Data is available upon request.

